# Intention of UK residents to wear facemasks and practise social distancing during the next respiratory virus pandemic

**DOI:** 10.64898/2026.05.21.26353824

**Authors:** David R M Smith, John Buckell, Thomas O Hancock, Liz Morrell, Koen B Pouwels

## Abstract

**Background:** Wearing facemasks and practising social distancing slow the spread of respiratory pathogens. However, in the event of a new pandemic emerging, the willingness of populations to voluntarily adopt these behaviours is unclear.

**Methods:** A discrete choice experiment was conducted among 2,006 UK-based adults. Participants were presented with hypothetical scenarios describing the emergence of a respiratory virus pandemic and were asked to choose when they would wear facemasks and practise social distancing. A mixed multinomial logit model was used to jointly estimate how disease severity and prevalence, uncertainty in these quantities, and individual-level characteristics influence behavioural choices.

**Findings:** Participants were averse to facemasks and social distancing in the absence of pandemic risk. For each ten-unit increase in severity (10 additional hospitalisations/1,000 infections), the odds of always wearing a facemask outside the home increased by 15.9% (95%CI: 14.3%, 17.5%), relative to rarely/never, and the odds of avoiding all people as much as possible increased by 16.4% (14.6%, 18.2%), relative to not avoiding anyone. Greater disease prevalence, uncertainty in disease severity or disease prevalence, a university education, prior COVID-19 vaccination and non-white ethnicity were also associated with choosing to always wear facemasks and avoid all people as much as possible. The probability of participants choosing to rarely/never wear facemasks varied from 13.4% (11.9%, 14.9%) in the lowest-risk scenario to 1.4% (1.2%, 1.7%) in the highest-risk scenario.

**Interpretation:** Perceived risks of disease and associated uncertainty drive intention of UK adults to adapt their behaviour in a future pandemic.

**Funding:** Medical Research Foundation.

**Research in Context:** *Evidence before this study:* In the aftermath of the COVID-19 pandemic, evidence on intended behaviour change during a future pandemic is scarce in the UK and globally. We searched PubMed with no language restrictions from 01/01/2020 to 18/05/2026 using the terms (“behavior” OR “behaviour” OR “mask” OR “distancing” OR “voluntary” OR “intention“) AND (“future pandemic” OR “next pandemic” OR “hypothetical pandemic” OR “Disease-X” OR “Pathogen-X”). An overview of UK biosecurity priorities highlights behaviour change as a critical yet understudied component of pandemic preparedness. A systematic review identified strong preferences for voluntary compliance with facemasks and social distancing, rather than mandatory enforcement. Choice experiments assessed the intention of different populations to get vaccinated or travel during the next pandemic, but no studies evaluated intention to wear facemasks and practise social distancing, nor associations between behaviour change and epidemic characteristics.

*Added value of this study:* This choice experiment among 2,006 UK-based adults is the first of its kind to assess population preferences for wearing facemasks and practising social distancing during the next respiratory virus pandemic. Although participants were averse to both behaviours in the absence of risk, they were increasingly likely to prefer them given higher estimates of disease prevalence or severity. Participants preferred to exercise greater caution when faced with greater epidemic uncertainty, and to exercise caution to a similar degree across both behaviours, instead of favouring one over the other. Associations with individual-level characteristics, including education, ethnicity and prior COVID-19 vaccination, highlight groups with less intention to modify their behaviour and potentially at greater risk of infection and transmission. These estimated relationships between behaviour change and epidemic characteristics may be harnessed in transmission dynamic modelling, outbreak forecasting and risk assessment.

*Implications of all the available evidence:* In the event of a future pandemic, adults in the UK intend to protect themselves and others to a greater degree when faced with increasing or more uncertain epidemic risk. Clear and concise communication of risk and its associated uncertainty may better enable populations to adapt their behaviour as appropriate to the evolving epidemiological context. Reducing uncertainty via investment in epidemiological surveillance could inadvertently reduce voluntary risk mitigation by excluding probabilities of very high risk.

## Introduction

Infectious disease transmission dynamics are shaped by how individuals choose to modify their behaviour, or not to, when faced with epidemic risk. Avoiding contact with others (*social distancing*) and wearing coverings over one’s nose and mouth (*facemask wearing*) are behaviours that can prevent catching and spreading airborne pathogens, and which are particularly important during emerging epidemics and in the absence of diagnostics, vaccines and therapeutics.^1,2^ During the COVID-19 pandemic, the enforcement of facemask wearing and social distancing via mandates and lockdowns is estimated to have saved many millions of lives globally.^3^ However, the social and economic costs of such measures, psychological reactions to the pandemic, political polarisation and other factors contributed to the development of staunch resistance to social distancing and facemask wearing among certain population groups.^4,5^

Behaviour change has been identified as a critical yet understudied pillar of UK biosecurity.^6^ A systematic review of population health preferences for behaviour change during infectious disease emergencies suggests that, overall, populations value facemask use and social distancing, with a clear preference for voluntary compliance over mandatory enforcement.^7^ Yet there is a lack of evidence on the willingness of individuals to voluntarily adopt infection prevention behaviours in the event of a new pandemic. Individuals behaving rationally may be expected to adapt their behaviour according to risk, with intention to wear facemasks and practise social distancing increasing with one’s perceived probability of falling seriously ill. However, the COVID-19 pandemic revealed extensive individual-level heterogeneity in the uptake of these behaviours, even during periods of heightened epidemic risk and prior to the availability of COVID-19 vaccines and antivirals.^8,9^ Now, in the aftermath of the COVID-19 pandemic, fatigue and scepticism towards transmission containment efforts could limit enthusiasm for voluntary behaviour change. Conversely, pandemic trauma or a desire to avoid “another COVID-19” could encourage individuals to adopt transmission prevention behaviours with zeal.

Anticipating voluntary behavioural responses is important for pandemic preparedness planning and risk communication.^10^ Understanding how individuals respond to perceived risk can help determine whether voluntary measures suffice or more restrictive measures may be necessary. Public behaviour during epidemics is shaped not only by perceived risk but also by how that risk is communicated.^11^ A better understanding of how perceived risk and uncertainty influence voluntary behaviour can therefore inform communication strategies and guide decisions about investing in early epidemiological studies to reduce uncertainty.

To understand how the UK population intends to behave during the early stages of the next respiratory virus pandemic, we conducted a discrete choice experiment (DCE) among a representative sample of adults living in the UK. We hypothesised that disease severity and prevalence, as well as uncertainty in these quantities and individual-level characteristics, influence the population’s intention to wear facemasks and practise social distancing.

## Methods

### Ethics

This study was approved by the University of Oxford’s Medical Sciences Interdivisional Research Ethics Committee (application ID 1080671).

### Discrete choice experiment (DCE)

A DCE is a preference elicitation method grounded in random utility theory.^12^ It is used to measure preferences by asking participants to choose between a set of alternatives presented in hypothetical scenarios (choice tasks). In each choice task, key characteristics (attributes) of the alternatives are varied, taking on different values (levels). As attribute levels are varied, participants are assumed to make choices that maximise their utility. Using choice modelling, DCE data can be analysed to estimate how attributes influence choices and associations with individual-level characteristics. In this study, a DCE administered as an online survey was used to understand preferences of UK residents for wearing facemasks and practising social distancing during a hypothetical respiratory virus pandemic. The structure and wording of the DCE – including choice alternatives, attributes and levels – were developed iteratively with multiple rounds of revision and editing to ensure clarity and consistency for a lay audience.

Participants were asked to describe their facemask wearing and social distancing practices over the month prior to the survey. Then, to establish the hypothetical pandemic context for this experiment, study participants were presented with the following scenario:

*A new infectious disease has recently been discovered and is spreading rapidly in humans. A global pandemic has just been declared by the World Health Organisation (WHO), and the disease is actively spreading throughout the UK. It has been identified as a viral infection of the respiratory tract, and people have started calling it Disease-X. However, this exact disease has never been seen before, and people have no pre-existing immunity to protect them from its effects. Scientists believe that it is mostly spread through the air when sick people cough or sneeze. No pandemic response measures have yet been imposed by the government, meaning that your response to this new disease is up to you*.

After reading this context, across 12 choice task study participants were asked to choose one of four alternatives for facemask wearing and one of four alternatives for social distancing. Finally, participants were asked to provide additional information about themselves.

To define appropriate alternatives, the UK government’s COVID-19 mandates, as well as quotes from experts published at the Science Media Centre in early 2020 with the tag COVID-19, were reviewed to evaluate wording and recommendations around these behaviours.^13,14^ The final list of alternatives is shown in **Table 1**. More details on the survey’s structure are described in the **Supplementary appendix** and the full survey text is provided online as a separate supplementary file.

**Table 1.** Choice alternatives (j) for the two included behaviours (b).

| Behaviour | Choice alternative |
| --- | --- |
| Facemask wearing<br>(b = M) | $j_0$ = rarely or never |
| | $j_1$ = only when around vulnerable people (e.g. in hospitals, nursing homes) |
| | $j_2$ = when in indoor public spaces (e.g. in shops, transport, cinema) |
| | $j_3$ = at all times outside of home |
| Social distancing<br>(b = D) | $j_0$ = don't actively avoid anyone |
| | $j_1$ = only avoid vulnerable people (e.g. the elderly, sick people) |
| | $j_2$ = avoid most people but still see friends and/or family |
| | $j_3$ = avoid all people as much as possible |

### Attributes and levels

An initial list of attributes and their levels was generated based on prior research on behavioural responses to infectious disease risk, including studies of risk perception, behavioural interventions, and epidemic dynamics. This list was refined through iterative discussion with a multidisciplinary team of infectious disease and DCE experts (n=12).

Perceived risk, a function of one’s perceived probability of acquiring infection and the perceived severity of outcomes conditional on infection, has been identified as a key determinant of behaviour change.^15^ The attribute *prevalence* was selected to capture risk of acquiring infection, with levels defined using prevalence estimates from waves of the COVID-19 pandemic.^16^ The attribute *severity* was selected to capture risk of consequences once infected, with levels defined using estimates of infection-fatality ratios of respiratory viruses responsible for major epidemics over recent years, including SARS-CoV-1 (2003), Influenza A(H1N1)pdm09 (2009) and SARS-CoV-2 (2020).^17–19^ Hospitalisation risk was identified as another key component of severity but, since hospitalisation risk and fatality risk are highly correlated, these were combined into a single severity attribute by assuming that the infection-fatality ratio is fixed at 25% of the infection-hospitalisation ratio in all scenarios.

The attributes *prevalence uncertainty* and *severity uncertainty* were selected, as prevalence and severity are often poorly quantified, particularly in the early stages of an epidemic, and uncertainty may impact confidence in reported estimates and, in turn, behaviour.^20,21^ Levels of uncertainty attributes were developed by simulating an epidemiological study measuring outcomes of interest, assuming limited sampling reflective of an early epidemic context, and assuming binomial outcome distributions to derive 95% confidence intervals (CIs). Estimates of low, medium and high uncertainty correspond, respectively, to 95% CIs derived from high (n=360), medium (n=80) and low (n=20) numbers of samples. Finally, the way in which epidemiological risk is communicated to the public can influence behaviour,^15^ so *risk communication* was selected as the final attribute. Experimental work suggests that visualising risk can decrease variance in risk interpretation,^22^ so the levels included were to present risk only using text *o*r using both text and graphical representations. The final list of attributes and levels is provided in **Table 2** and an example choice task is provided in **Figure 1**.

**Figure 1.**
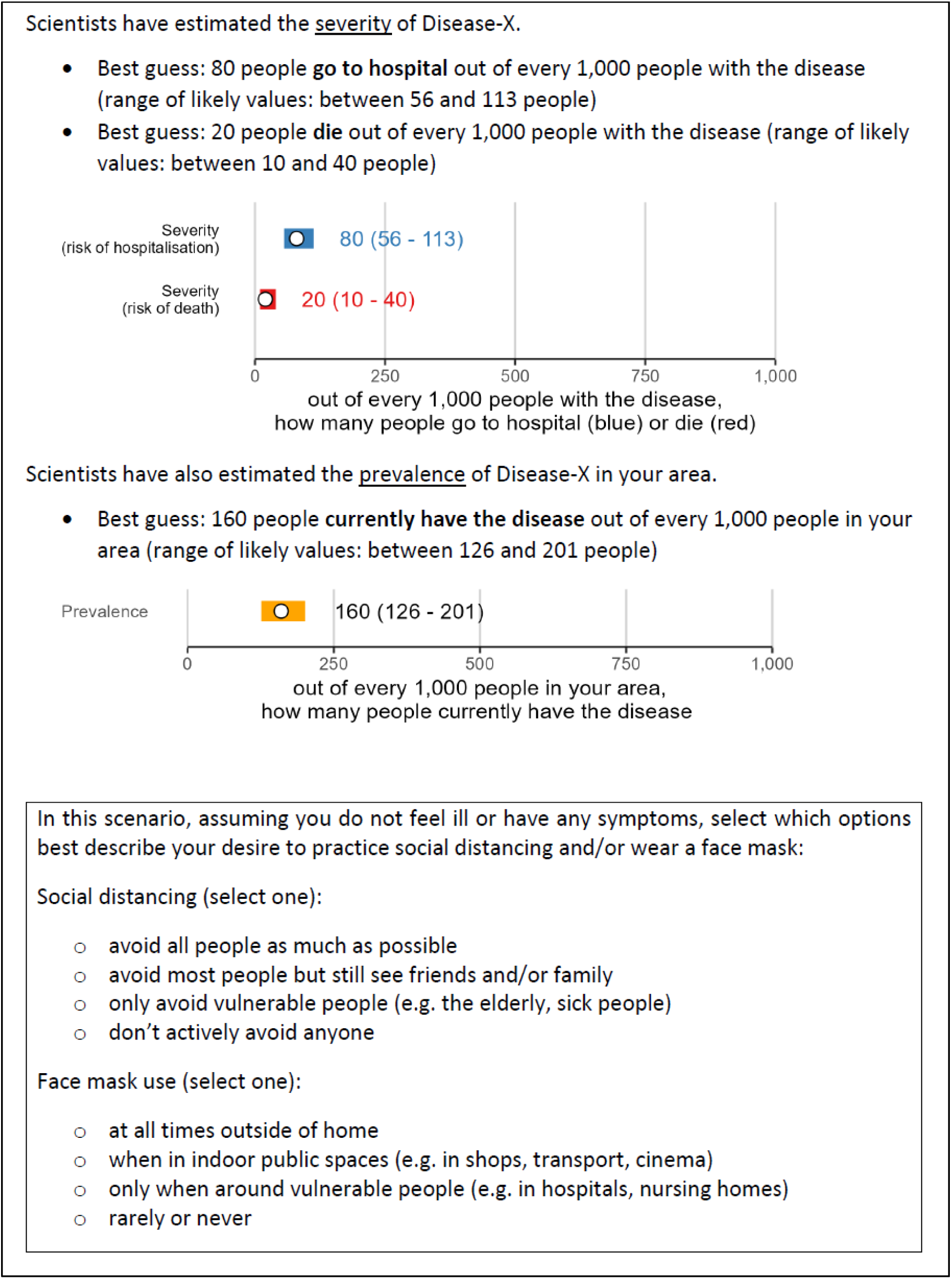
Example of a choice task from the survey. Attribute levels of this task are: moderate severity, low severity uncertainty, high prevalence, low prevalence uncertainty, and risk communication including both text and visual.

**Table 2.**
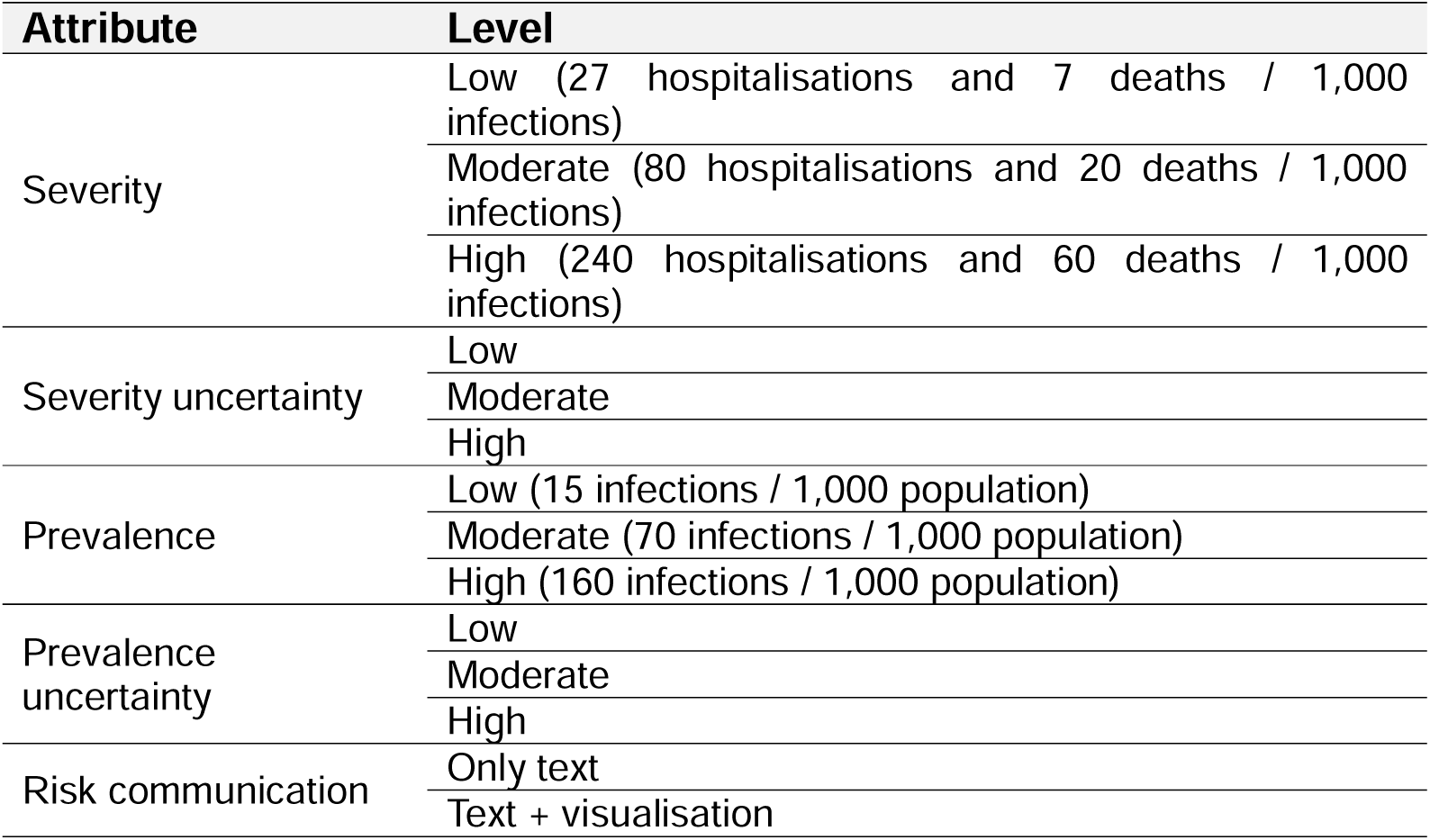
Attributes and levels. The specific quantities corresponding to each level of severity uncertainty and prevalence uncertainty depend on corresponding levels of severity and prevalence, respectively (see **Table S1** for a complete list and **Figure 1** for an example).

| Attribute | Level |
| --- | --- |
| Severity | Low (27 hospitalisations and 7 deaths / 1,000 infections) |
|  | Moderate (80 hospitalisations and 20 deaths / 1,000 infections) |
|  | High (240 hospitalisations and 60 deaths / 1,000 infections) |
| Severity uncertainty | Low |
|  | Moderate |
|  | High |
| Prevalence | Low (15 infections / 1,000 population) |
|  | Moderate (70 infections / 1,000 population) |
|  | High (160 infections / 1,000 population) |
| Prevalence uncertainty | Low |
|  | Moderate |
|  | High |
| Risk communication | Only text |
|  | Text + visualisation |

### Experimental design and survey administration

A full factorial survey design including all possible combinations of included attributes and levels would lead to an infeasibly long survey. An efficient DCE design refers to the combination of attributes and levels that optimises parameter identification and statistical efficiency. A pilot DCE was developed first to generate informative Bayesian priors for the final design. D-optimal designs with 12 choice tasks were identified for both the pilot and final designs by specifying a multinomial logit (MNL) model in Ngene software (version 1.4.0). All participants received the same survey, but several components were randomised to prevent anchoring or question order bias. See the **Supplementary appendix** and **Tables S2, S3 and S4** for more information.

The pilot and final surveys were available online only and administered by Dynata, a survey company and online panel provider (https://www.dynata.com/). Standard data quality checks were applied. The pilot survey was launched on 8 April 2025 and 51 complete survey responses were obtained. The final survey was launched on 19 May 2025 and ran until 10 June 2025. Dynata recruited participants using their online panels, limiting recruitment to UK residents aged 18+ with no other exclusion criteria. For the final survey, quotas were set to ensure the survey sample was representative of adults residing in the UK in terms of age, gender and region of residence (**Table S5**). Pilot responses were not included in analysis of the final survey due to differences in wording and design.

### Statistical analysis

For the primary analysis, a mixed multinomial logit model was specified with an indirect utility function comprising deterministic and stochastic components, in which the utilities associated with facemask wearing choices and social distancing choices were estimated jointly. To account for correlation between these choices within each choice task, alternative-specific individual-level random effects were included, capturing latent affinity for specific alternatives and inducing utility correlation across choices. This model is given by,

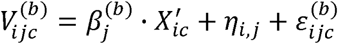

where 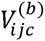 is the indirect utility of individual *i* from alternative *j* in choice task *c* for behaviour *b*; 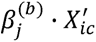 is the deterministic utility component comprising included, attributes, 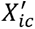, and utility weights (or preferences) for those attributes, 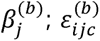 is the random component of utility; and 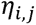 is the random latent affinity of individual *i* for alternative *j*,

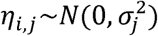

where this term is shared across the jth alternative of both choice behaviours and therefore captures to what degree individuals make consistent choices across behaviours within each choice task (for example, choosing *j*=2 for both facemask wearing and social distancing). A stronger correlation implies individuals are more likely to choose facemask and social distancing behaviours at the same jth level. Correlations were not considered between asymmetric choice levels (e.g. j=1 for facemasks use and j=3 for social distancing), following strong correlation observed only across symmetric jth alternatives in survey responses (**Figure S1**). Additional details on model specification are provided in the **Supplementary appendix**.

Deterministic preference heterogeneity was captured by incorporating individual-level participant characteristics as covariates. To avoid over-parameterisation and unstable estimation due to sparse categories, levels of categorical variables were collapsed where appropriate to ensure sufficient observations within each category (**Table S6**). Covariates were selected following iterative rounds of forwards-backwards stepwise model selection and interaction testing guided by the Bayesian Information Criteria (BIC), balancing improvements in fit against model complexity (see **Table S7**). For participant age, the only integer variable, binning individuals into age tertiles led to best fit of the functional forms tested (linear, log-linear, quadratic, cubic, tertiles, quartiles).

Model estimation was conducted in R (version 4.4.2) using the package *Apollo* (version 0.3.5), with simulated maximum likelihood estimation implemented using 500 Halton draws to approximate the integrals over the distribution of random parameters. Statistical significance was assessed using robust standard errors and robust two-tailed t-ratios, and main results are presented as adjusted odds ratios (ORs) and 95% confidence intervals (95%CI). For ease of interpretation, ORs for severity and prevalence, and any interactions with them, were rescaled by multiplying coefficients and robust standard errors by a factor of ten, such that their interpretation is the odds of choosing an alternative *j* relative to the reference (*j*=0) given a ten-point increase in severity or prevalence, equivalent to a single percentage point increase.

After model estimation, choice probability predictions were generated for the full survey population across a range of scenarios. Choice probabilities were derived from estimated model parameters using Krinsky–Robb draws (n=250), whereby repeated samples were taken from the asymptotic multivariate normal distribution of the estimated coefficients to propagate parameter uncertainty into predicted outcomes. For the attributes interpreted as continuous variables (severity and prevalence), choice behaviours were predicted over a range of 10 values, uniformly distributed from the lowest to highest levels included in the DCE. Final predictions are presented as means and the 2.5^th^ and 97.5^th^ quantiles, i.e. 95% uncertainty intervals (95%UI) of the predicted outcome distribution. Unless otherwise specified, the reference case against which attributes are varied is one of least risk: low estimates of severity and prevalence, low severity uncertainty and prevalence uncertainty, and the communication of risk including both text and visualisation.

Sensitivity analyses were conducted to assess robustness of findings. First, participants responding “prefer not to answer” to any question (approximately 8% of the total sample) were removed and the primary model was re-estimated. Second, the variables describing participants’ facemask wearing and social distancing choices over the month prior to the survey were removed, resulting in selection and estimation of an alternative model including only individual-level characteristics as covariates, and not individual-level choices, making results more easily generalisable (**Table S8**).

## Results

### Participant characteristics

A complete survey response was received from n=2,008 study participants. Two responses were removed due to data quality issues, for a final sample of n=2,006 participants. The median age was 49 years [IQR: 33, 64], 51.5% (n=1,034) were female and 84.4% (n=1,694) resided in England (**Table 3**). Four out of five participants reported white ethnicity (n=1,596, 79.6%), while roughly half reported living in a city (n=1,034; 51.5%), having a university degree or equivalent (n=991; 49.4%), or having annual household income exceeding £40,000 (n=977, 48.7%). One in seven participants reported suffering from a serious chronic illness (n=282, 14.1%) or living with or caring for any fragile or vulnerable people (n=287, 14.3%). Over the month prior to the survey, 61.6% of participants (n=1,236) rarely or never wore a facemask and 55.0% (n=1,104) didn’t actively avoid anyone. Conversely, 11.9% (n=238) wore a facemask at all times outside the home and 11.2% (n=225) avoided all people as much as possible (**Figure S2**). Written survey feedback was largely positive and the majority of participants found the survey questions easy to understand and the pictures provided helpful (**Figure S3**).

**Table 3.**
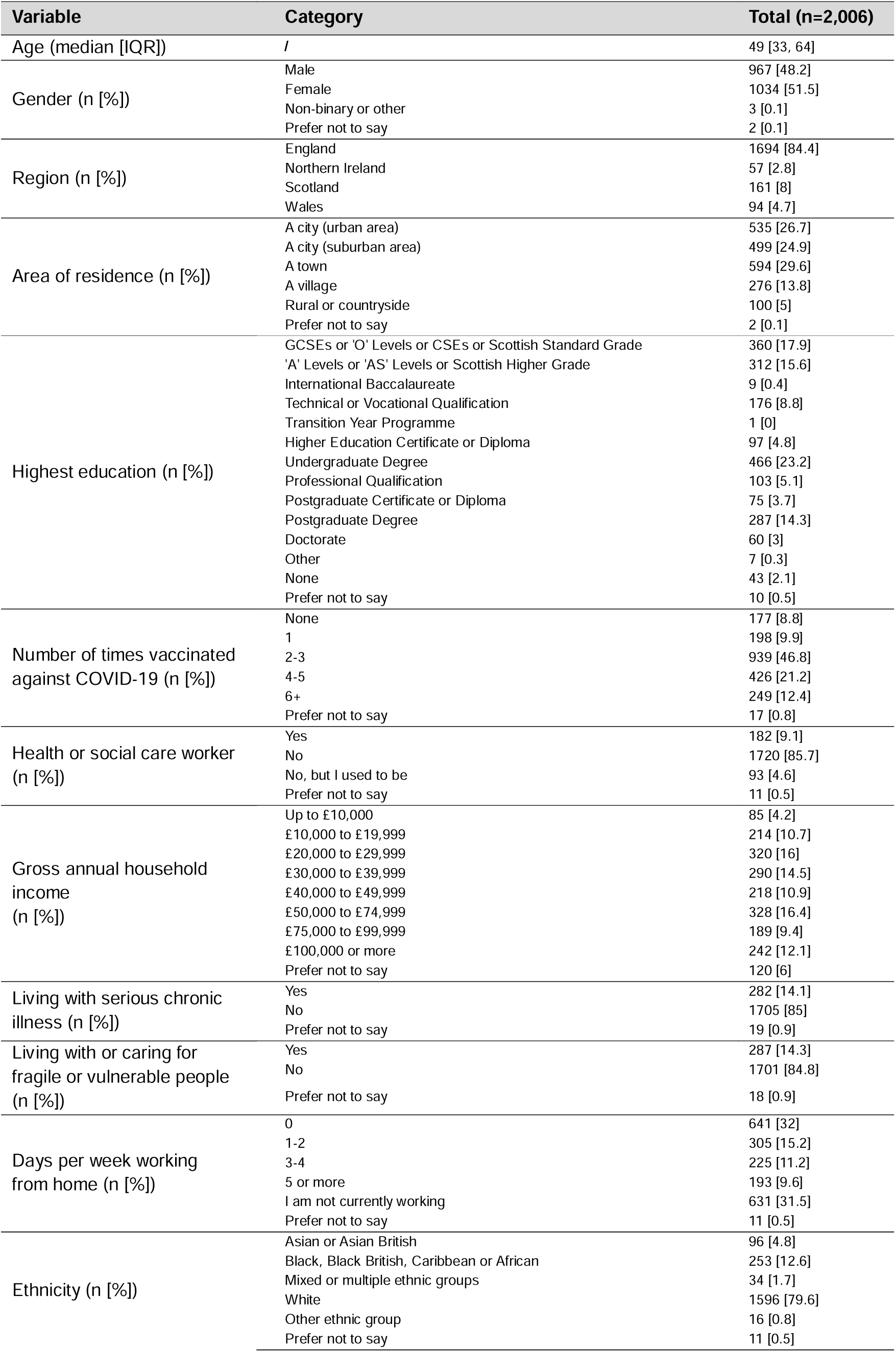
Participant characteristics.

### Main effects

Disease severity was strongly associated with increased facemask wearing and social distancing (**Table 4**; see **Table S9** for coefficients). Relative to rarely/never wearing a facemask (*j*=0), each ten-unit increase in severity (10 additional hospitalisations and 2.5 additional deaths/1,000 infections) was associated with, respectively, 4.1% (95%CI: 0.3%, 5.3%), 9.9% (8.5%, 11.3%), and 15.9% (14.3%, 17.5%) greater odds of choosing to wear facemasks only around vulnerable people (*j*=1), in indoor public spaces (*j*=2), and at all times outside the home (*j*=3). For social distancing, relative to not actively avoiding anyone (*j*=0), each ten-unit increase in severity was associated with, respectively, 3.5% (2.3%, 4.7%), 9.4% (7.9%, 10.9%), and 16.4% (14.6%, 18.2%) greater odds of avoiding only vulnerable people (*j*=1), most people other than family/friends (*j*=2), and all people as much as possible (*j*=3).

**Table 4.**
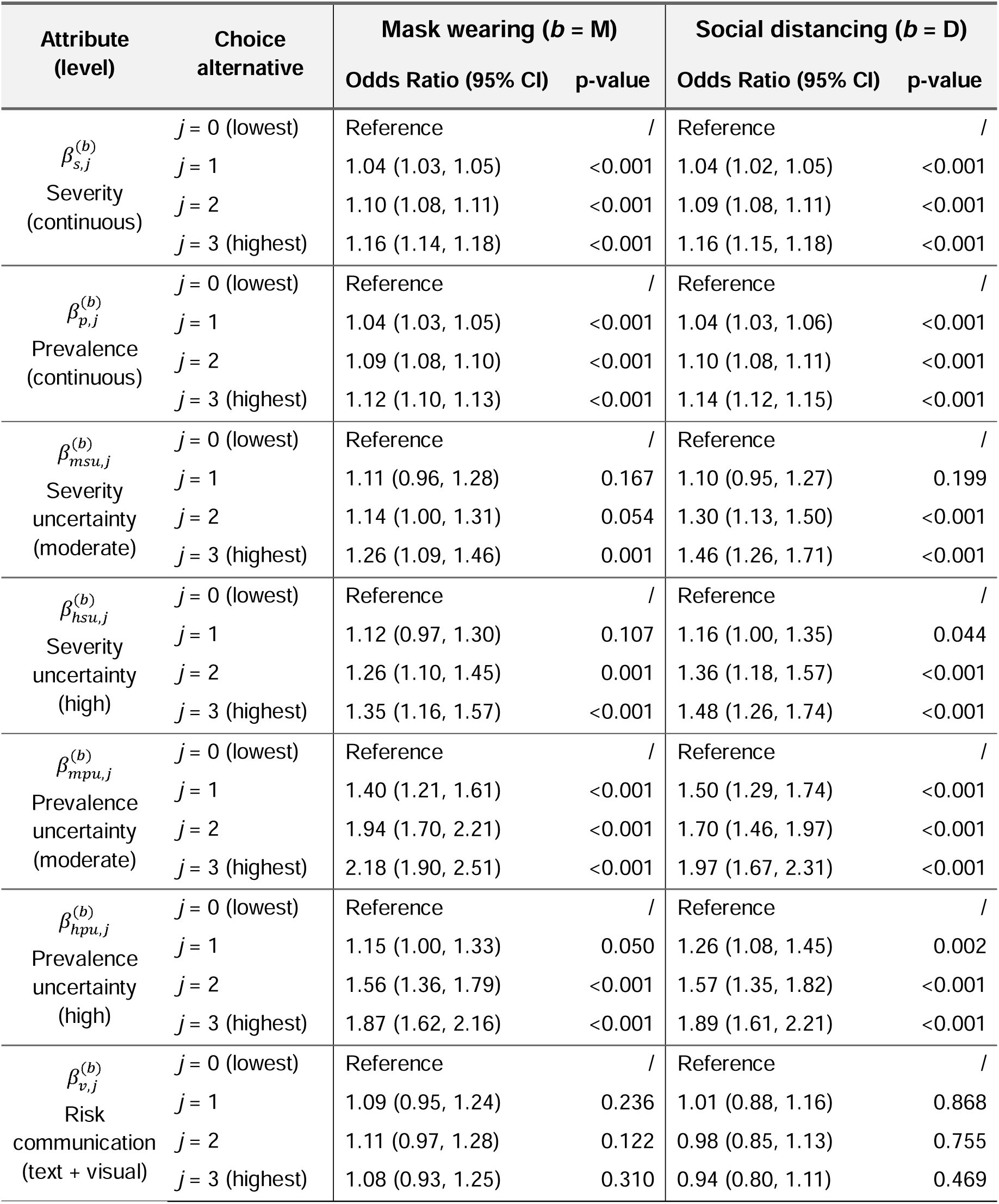
Main effects. Model estimates from the joint multinomial logit model, presented as Odds Ratios and two-tailed p-values (α=0.05). Choice alternatives for facemask wearing range from rarely/never (j=0) to all times outside the home (j=3) and for social distancing range from not actively avoiding anyone (j=0) to avoiding all people as much as possible (j=3) (see Table 1). These effects were estimated in a model adjusting for deterministic preference heterogeneity in the form of individual-level covariates (see Table 5). Odds Ratios for severity and prevalence are choices interpreted as corresponding to each ten-point increase in severity (10 additional hospitalisations and 2.5 additional deaths per 1,000 infections) and prevalence (infections/1,000 population).

| Attribute<br>(level) | Choice<br>alternative | Mask wearing ($b = M$) | | Social distancing ($b = D$) | |
| --- | --- | --- | --- | --- | --- |
|  |  | Odds Ratio (95% CI) | p-value | Odds Ratio (95% CI) | p-value |
| $\beta_{s,j}^{(b)}$<br>Severity<br>(continuous) | $j = 0$ (lowest) | Reference | / | Reference | / |
| | $j = 1$ | 1.04 (1.03, 1.05) | <0.001 | 1.04 (1.02, 1.05) | <0.001 |
| | $j = 2$ | 1.10 (1.08, 1.11) | <0.001 | 1.09 (1.08, 1.11) | <0.001 |
| | $j = 3$ (highest) | 1.16 (1.14, 1.18) | <0.001 | 1.16 (1.15, 1.18) | <0.001 |
| $\beta_{p,j}^{(b)}$<br>Prevalence<br>(continuous) | $j = 0$ (lowest) | Reference | / | Reference | / |
| | $j = 1$ | 1.04 (1.03, 1.05) | <0.001 | 1.04 (1.03, 1.06) | <0.001 |
| | $j = 2$ | 1.09 (1.08, 1.10) | <0.001 | 1.10 (1.08, 1.11) | <0.001 |
| | $j = 3$ (highest) | 1.12 (1.10, 1.13) | <0.001 | 1.14 (1.12, 1.15) | <0.001 |
| $\beta_{msu,j}^{(b)}$<br>Severity<br>uncertainty<br>(moderate) | $j = 0$ (lowest) | Reference | / | Reference | / |
| | $j = 1$ | 1.11 (0.96, 1.28) | 0.167 | 1.10 (0.95, 1.27) | 0.199 |
| | $j = 2$ | 1.14 (1.00, 1.31) | 0.054 | 1.30 (1.13, 1.50) | <0.001 |
| | $j = 3$ (highest) | 1.26 (1.09, 1.46) | 0.001 | 1.46 (1.26, 1.71) | <0.001 |
| $\beta_{hsu,j}^{(b)}$<br>Severity<br>uncertainty<br>(high) | $j = 0$ (lowest) | Reference | / | Reference | / |
| | $j = 1$ | 1.12 (0.97, 1.30) | 0.107 | 1.16 (1.00, 1.35) | 0.044 |
| | $j = 2$ | 1.26 (1.10, 1.45) | 0.001 | 1.36 (1.18, 1.57) | <0.001 |
| | $j = 3$ (highest) | 1.35 (1.16, 1.57) | <0.001 | 1.48 (1.26, 1.74) | <0.001 |
| $\beta_{mpu,j}^{(b)}$<br>Prevalence<br>uncertainty<br>(moderate) | $j = 0$ (lowest) | Reference | / | Reference | / |
| | $j = 1$ | 1.40 (1.21, 1.61) | <0.001 | 1.50 (1.29, 1.74) | <0.001 |
| | $j = 2$ | 1.94 (1.70, 2.21) | <0.001 | 1.70 (1.46, 1.97) | <0.001 |
| | $j = 3$ (highest) | 2.18 (1.90, 2.51) | <0.001 | 1.97 (1.67, 2.31) | <0.001 |
| $\beta_{hpu,j}^{(b)}$<br>Prevalence<br>uncertainty<br>(high) | $j = 0$ (lowest) | Reference | / | Reference | / |
| | $j = 1$ | 1.15 (1.00, 1.33) | 0.050 | 1.26 (1.08, 1.45) | 0.002 |
| | $j = 2$ | 1.56 (1.36, 1.79) | <0.001 | 1.57 (1.35, 1.82) | <0.001 |
| | $j = 3$ (highest) | 1.87 (1.62, 2.16) | <0.001 | 1.89 (1.61, 2.21) | <0.001 |
| $\beta_{v,j}^{(b)}$<br>Risk<br>communication<br>(text + visual) | $j = 0$ (lowest) | Reference | / | Reference | / |
| | $j = 1$ | 1.09 (0.95, 1.24) | 0.236 | 1.01 (0.88, 1.16) | 0.868 |
| | $j = 2$ | 1.11 (0.97, 1.28) | 0.122 | 0.98 (0.85, 1.13) | 0.755 |
| | $j = 3$ (highest) | 1.08 (0.93, 1.25) | 0.310 | 0.94 (0.80, 1.11) | 0.469 |

Disease prevalence was also strongly associated with facemask wearing and social distancing. Relative to rarely/never wearing a facemask, each ten-unit increase in prevalence (10 additional infections/1,000 population in the local area) was associated with, respectively, 4.1% (95%CI: 2.9%,5.3%), 8.7% (95%CI: 7.5%, 9.9%), and 11.8% (95%CI: 10.5%, 13.2%) greater odds of choosing to wear a facemask only around vulnerable people, in indoor public spaces, and at all times outside the home. Relative to not actively avoiding anyone, each ten-unit increase in prevalence was associated with, respectively, 4.4% (3.2%, 5.6%), 9.6% (8.3%, 10.9%), and 13.8% (12.3%, 15.3%) greater odds of avoiding only vulnerable people, most people other than family/friends, and all people as much as possible.

Moderate or high levels of epidemiological uncertainty, which imply higher ranges of possible outcomes, were generally positively associated with increased facemask wearing and social distancing. Given high uncertainty in true disease severity, participants had 35% (95%CI: 16%, 57%) greater odds of choosing to wear a facemask at all times outside the home and 48% (95%CI: 26%, 74%) greater odds of choosing to avoid all people as much as possible. However, when presenting estimates of disease prevalence and severity to participants, including a figure (bar plot) in addition to text had no impact on their choices. Main effect estimates were largely consistent in sensitivity analyses (**Figure S4**).

### Participant heterogeneity

Several individual-level covariates were associated with preferences for facemask wearing and social distancing (**Table 5**; see **Tables S10, S11** for coefficients). At baseline, intercept terms suggest that participants were overall averse to facemask wearing and social distancing (**Table S10**). Practising cautious behaviour (any amount of facemask wearing or social distancing) over the month prior to the survey was strongly associated with choosing to wear a facemask or practise social distancing during the next pandemic. An interaction between this covariate and severity was negatively associated with greater facemask wearing and social distancing, suggesting that impacts of severity on choices are greater among those not already practising infection prevention behaviours. Prior COVID-19 vaccination was strongly associated with both behaviours, especially among participants having received 4 or more doses. Non-white ethnicity and having a university degree or equivalent were both associated with highest levels of facemask wearing and distancing. Individual-level error correlation terms were also highly significant (**Table S11**), suggesting that individuals are consistent in their degree of adherence to both facemask use and social distancing behaviours, particularly among those choosing to practise both behaviours as much as possible (j=3).

**Table 5.** Deterministic heterogeneity. Impacts of covariates included in the joint multinomial logit model on participant preferences, presented as Odds Ratios and two-tailed p-values (α=0.05). Choice alternatives for facemask wearing range from rarely/never (j=0) to all times outside the home (j=3) and for social distancing range from not actively avoiding anyone (j=0) to avoiding all people as much as possible (j=3) (see Table 1). Main effects from the model corresponding with these estimates are presented in Table 4.

| Covariate | Choice alternative | Facemask wearing ($b = M$) | | Social distancing ($b = D$) | |
| --- | --- | --- | --- | --- | --- |
|  |  | Odds Ratio (95% CI) | p-value | Odds Ratio (95% CI) | p-value |
| $\beta_{basebehav,j}^{(b)}$<br>Cautious current behaviour | $j = 0$ (lowest) | Reference | / | Reference | / |
| | $j = 1$ | 4.94 (2.88, 8.46) | <0.001 | 4.02 (2.41, 6.69) | <0.001 |
| | $j = 2$ | 6.60 (3.76, 11.59) | <0.001 | 6.01 (3.62, 9.97) | <0.001 |
| | $j = 3$ (highest) | 17.39 (9.59, 31.55) | <0.001 | 11.75 (6.67, 20.73) | <0.001 |
| $\beta_{education,j}^{(b)}$<br>Higher education | $j = 0$ (lowest) | Reference | / | Reference | / |
| | $j = 1$ | 1.2 (0.70, 2.08) | 0.505 | 1.77 (1.03, 3.04) | 0.040 |
| | $j = 2$ | 1.27 (0.76, 2.12) | 0.353 | 1.47 (0.92, 2.37) | 0.110 |
| | $j = 3$ (highest) | 1.97 (1.05, 3.70) | 0.035 | 2.39 (1.31, 4.37) | 0.005 |
| $\beta_{vaxfew,j}^{(b)}$<br>1 to 3 COVID-19 vaccine doses | $j = 0$ (lowest) | Reference | / | Reference | / |
| | $j = 1$ | 3.45 (1.86, 6.40) | <0.001 | 2.13 (1.17, 3.89) | 0.013 |
| | $j = 2$ | 10.20 (4.46, 23.32) | <0.001 | 4.52 (2.05, 9.96) | <0.001 |
| | $j = 3$ (highest) | 5.11 (2.33, 11.20) | <0.001 | 3.63 (1.64, 8.04) | 0.001 |
| $\beta_{vaxmany,j}^{(b)}$<br>4+ COVID-19 vaccine doses | $j = 0$ (lowest) | Reference | / | Reference | / |
| | $j = 1$ | 3.78 (1.80, 7.96) | <0.001 | 1.75 (0.86, 3.55) | 0.124 |
| | $j = 2$ | 21.92 (9.33, 51.49) | <0.001 | 7.72 (3.46, 17.22) | <0.001 |
| | $j = 3$ (highest) | 21.87 (9.76, 49.00) | <0.001 | 12.68 (5.76, 27.91) | <0.001 |
| $\beta_{white,j}^{(b)}$<br>White ethnicity | $j = 0$ (lowest) | Reference | / | Reference | / |
| | $j = 1$ | 0.74 (0.38, 1.47) | 0.390 | 1.07 (0.56, 2.04) | 0.843 |
| | $j = 2$ | 0.85 (0.43, 1.68) | 0.640 | 1.01 (0.54, 1.89) | 0.982 |
| | $j = 3$ (highest) | 0.18 (0.08, 0.39) | <0.001 | 0.41 (0.19, 0.88) | 0.022 |
| $\gamma_{sev,basebehav}^{(b)}$<br>Current behaviour and severity interaction | $j = 0$ (lowest) | Reference | / | Reference | / |
| | $j = 1$ | 0.99 (0.96, 1.01) | 0.244 | 0.98 (0.96, 1.00) | 0.101 |
| | $j = 2$ | 0.96 (0.94, 0.98) | 0.002 | 0.96 (0.93, 0.98) | <0.001 |
| | $j = 3$ (highest) | 0.94 (0.91, 0.96) | <0.001 | 0.93 (0.91, 0.95) | <0.001 |

### Predicted behaviour

Predicted behaviours were calculated by averaging individual-level predicted choice probabilities across the study sample, producing sample-averaged estimates under different scenarios. Probabilities of wearing facemasks in indoor public spaces or at all times outside the home increased with severity and prevalence, offset by declines in probabilities of wearing facemasks only when around vulnerable people or rarely/never (**Figure 2**). Similarly, probabilities of avoiding all people or maintaining contact only with family/friends increased with severity and prevalence, offset by declines in probabilities of only avoiding vulnerable people or not actively avoiding anyone (**Figure 2**). In sensitivity analyses, some individual-level covariates and their estimates varied (**Figure S5**), but choice predictions were nonetheless consistent with the primary analysis (**Figures S6, S7**).

**Figure 2.**
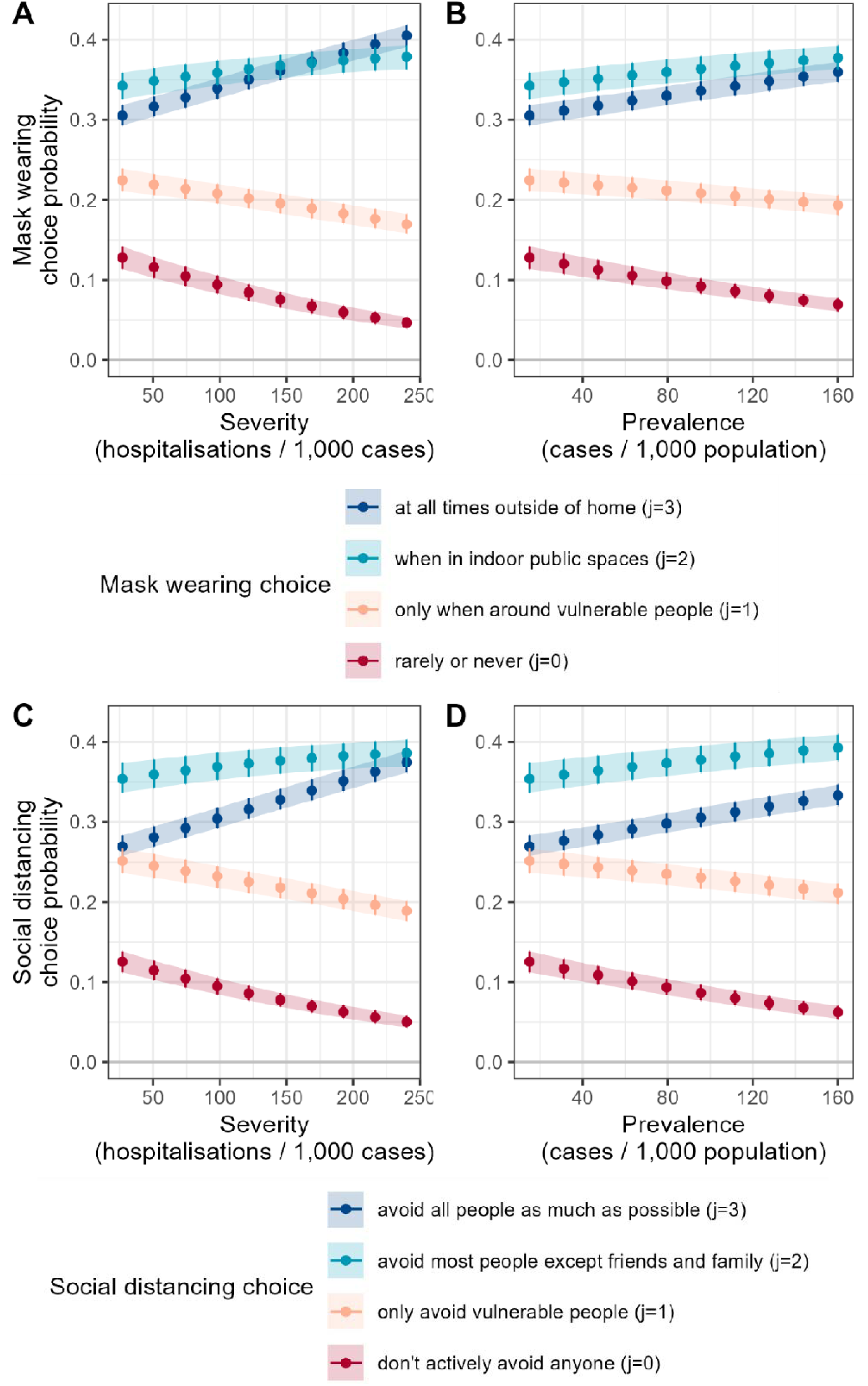
Predicted probabilities of behavioural choices among the sample population when varying disease severity and prevalence from the lowest to highest considered levels. All predictions assume low severity uncertainty, low prevalence uncertainty and the communication of risk including visualisation. When varying severity (panels **A**, **C**) low prevalence is assumed, and when varying prevalence (panels **B**, **D**) low severity is assumed. Points and error bars represent, respectively, means and 95% bootstrap prediction intervals (n=250).

In the lowest-risk scenario including low uncertainty, this population was more likely to wear facemasks only in indoor public spaces (30.5%; 95%UI: 29.3%, 31.8%) than at all times outside of home (33.8%; 32.2%, 35.3%), and more likely to maintain contacts with family/friends (35.4%; 33.8%, 37.2%) than to avoid all people as much as possible (27.3%; 26.0%, 28.7%). Probabilities of choosing to never/rarely wear facemasks (13.4%; 11.9%, 14.9%) and to not avoid anyone in particular (12.4%; 11.0%, 14.0%) were similar. By contrast, in the highest risk scenario including high uncertainty, the dominant choices were to wear a facemask at all times outside the home (48.2%; 46.7%, 49.8%) and avoid all people as much as possible (46.6%; 45.2%, 48.1%), with low probabilities of choosing to rarely/never wear a facemask (1.4%; 1.2%, 1.7%) or not actively avoid anyone (1.1%; 0.9%, 1.4%). Variation in severity uncertainty and prevalence uncertainty had comparatively less impact on predicted behaviour than variation in severity and prevalence (**Figures S8, S9**).

Compared to the low-risk reference case, **Figure 3** shows how probabilities of different behaviours changed given changes in attribute levels. High severity was the attribute level leading to greatest change, including increases of 0.101 (0.093, 0.109) in the probability of wearing a facemask at all times outside of home and of 0.105 (0.098, 0.113) in the probability of avoiding all people as much as possible.

**Figure 3.**
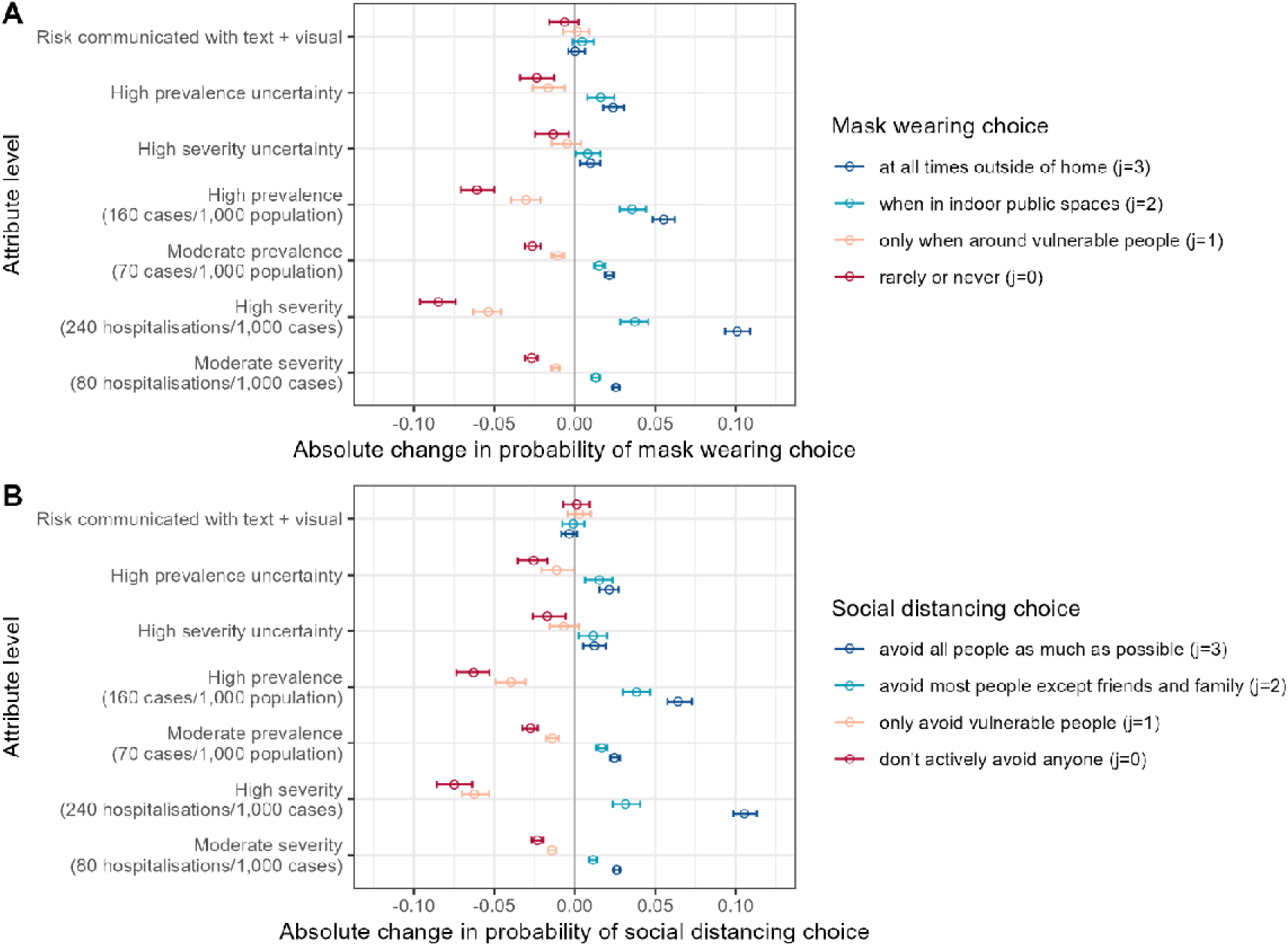
Effects of changing attribute levels on predicted probabilities of facemask wearing choices (**A**) and social distancing choices (**B**). The reference case here is: low severity (27 hospitalisations/1,000 cases and 7 deaths/1,000 cases), low prevalence (15 cases/1,000 population), low severity uncertainty, low prevalence uncertainty and the communication of risk only including text. See **Table S1** for quantities corresponding with different levels of prevalence and severity uncertainty. Points and error bars represent, respectively, means and 95% bootstrap prediction intervals (n=250).

## Discussion

This study sought to measure the intention of the UK population to voluntarily wear facemasks and practise social distancing in the early stages of the next respiratory virus pandemic. Our findings suggest that UK adults are averse to these behaviours in the absence of risk but are increasingly likely to practise them with increasing disease prevalence or severity. After declaration of a (hypothetical) pandemic by the WHO, in a low risk scenario (local prevalence = 1.5%, infection fatality rate <1%, low uncertainty), there was substantial heterogeneity in this population’s intention to wear facemasks [at all times outside the home (30.5%); in indoor public spaces (33.8%); only around vulnerable people (22.3%); rarely/never (13.4%)] and practise social distancing [avoid all people as much as possible (27.2%); avoid most people except friends/family (35.5%); avoid only vulnerable people (24.8%); avoid no one in particular (12.4%)]. By contrast, in a high risk scenario (local prevalence = 16%, infection fatality rate = 6%, high uncertainty), there was convergence towards substantial facemask use [at all times outside the home (48.2%); in indoor public spaces (39.2%); only around vulnerable people (11.2%); rarely/never (1.4%)] and social distancing [avoid all people (46.6%); avoid most people except friends/family (40.2%); avoid only vulnerable people (11.4%); avoid no one in particular (1.1%)]. These results highlight an overall rational response to a future pandemic, with individuals choosing to protect themselves and others to a greater degree when faced with increasing risk.

Evidence of strong correlation between facemask and social distancing choices at the individual-level further suggests that individuals prefer to exercise caution to a similar degree across multiple behaviours. For example, individuals who preferred to wear facemasks only around vulnerable people also preferred to only distance themselves from vulnerable people, while individuals who preferred to wear facemasks at all times outside the home also preferred to avoid all people as much as possible. Therefore, these results do not support the hypothesis of risk compensation, i.e. that individuals may be comfortable not socially distancing in the early stages of the next pandemic because they could instead wear a facemask, or vice versa.

Comparable evidence on intended behaviour during a future pandemic is scarce in the UK and globally. Nonetheless, behavioural intention in the present work can be compared, with caution, to voluntary behaviours reported during the COVID-19 pandemic. In surveys from England and Wales conducted during a period in 2021 with no legally enforced COVID-19 restrictions, 88.5% of respondents reported wearing facemasks in indoor public places and 86.8% reported maintaining social distancing all or most of the time.^23^ In autumn 2020 in British Columbia, Canada, prior to the enforcement of a provincial facemask mandate, roughly 80% of respondents reported any use of facemasks.^24^ Broadly speaking, these figures are similar to our estimates of intended behaviours in higher-risk pandemic scenarios.

Our findings also highlight that clear and concise communication of uncertainty in risk may better enable populations to adapt their behaviour. When uncertainty in prevalence and severity estimates increased, so too did individuals’ propensity to choose to adopt infection prevention behaviours. This suggests a desire to exercise more caution in more uncertain conditions. Previous experimental studies have identified relatively minor effects of communicating uncertainty or not on public trust and source reliability.^25,26^ However, impacts of uncertainty communication on behaviour change have been less well understood, particularly in rapidly evolving and highly uncertain early epidemic contexts.

Adjusting for individual-level covariates revealed associations of current cautious behaviour, prior COVID-19 vaccination, higher educational attainment and non-white ethnicity with choosing to wear facemasks and practise social distancing. These associations should be interpreted only as descriptive of preference heterogeneity in this sample. Previous UK studies have reported mixed findings regarding sociodemographic predictors of facemask use and social distancing during the COVID-19 pandemic. A survey of adults from May 2020 found associations of female gender, but not age, education or urbanicity with adherence to lockdown measures;^27^ and household surveys from 2021 found associations of older age and female gender with social distancing and facemask use, and of non-white ethnicity with social distancing but not facemask use.^23^

Findings from this study may be used to better anticipate and predict human behaviour during future emerging infectious disease epidemics. Due to data limitations, epidemiological models rarely account for time-varying behaviour change, even as new risk estimates become available and populations adapt. Incorporating behavioural adaptation into epidemiological models using endogenous feedback loops that account for dynamic associations between measures of risk (infection prevalence, disease severity, uncertainty in these measures) and behaviour could lead to more accurate forecasting of disease spread. Furthermore, our results suggest that transparent communication of epidemiological uncertainty may not only be ethical but also effective for disease control, prompting individuals to exercise more caution given greater uncertainty in true risk. Conversely, policy makers should consider that reducing epidemiological uncertainty via investment in large-scale studies such as the COVID-19 Infection Survey could inadvertently reduce voluntary risk mitigation by excluding probabilities of very high risk. This does not diminish the importance of rigorous surveillance systems to inform public policy, but highlights the need to anticipate how evolving risk estimates, and their communication, may shape population behaviour over time.

Strengths of our study include use of mixed effects modelling accounting for individual-level random preferences and correlation between facemask and social distancing choices, as well as our finding of consistent probability predictions across included sensitivity analyses. Most participants reported finding the survey questions easy or very easy to understand and found the visuals to be helpful when responding, despite the inclusion of visuals being found to have no impact on choices. The latter result is consistent with an experimental finding that including a pictograph of risk in addition to a percentage estimate had no impact on risk interpretation.^28^ That study did, however, find a strong impact of qualitative statements assessing evidence quality, which was not explored in the present work. Future research should examine how communicated uncertainty interacts with source credibility to influence behavioural responses. Such effects may operate through mechanisms distinct from statistical uncertainty as explored here, including trust, social identity, and prior beliefs. The communication platform, e.g. television, radio, social media or word-of-mouth, may further shape how risk information is interpreted and acted upon.

This work has several limitations. As in all stated choice experiments, the provided survey context was hypothetical, meaning choices were made without the stresses of a real-world situation. However, all participants recently lived through the COVID-19 pandemic and may have drawn upon that experience to inform their responses. Unlike in a real-world scenario, all recipients also received their information from the same source, while in reality members of the general public receive their information from heterogeneous sources across diverse platforms, and disinformation and misinformation may play an important role in risk perception and behaviour during a future pandemic, which we could not account for here. Finally, despite the use of quotas for age, sex, and region of residence, our sample is not fully representative of the UK population. The reported median gross household income (roughly £40,000) was comparable to official UK estimates for 2024 (£44,700);^29^ and reported white ethnicity (80%) was comparable to 2021 census estimates for England and Wales (82%).^30^ However, the share of our sample having a university degree or equivalent (49%) exceeded census estimates for England and Wales (34%). This suggests potential overestimation of facemask use and social distancing, given our finding of an association between higher education and these behaviours. Further, relatively high shares of the survey population responded that, over the month prior to the survey, they wore a mask at all times outside the home (12%) or avoided all people as much as possible (11%). While motivations for these behaviours could be numerous – including to prevent catching infection, protect loved ones, avoid allergens, manage social anxiety, and respect socio-cultural and religious norms – it is possible that some participants were subject to social desirability bias (that is, being motivated to make choices that put them in a positive light).

In conclusion, this study provides, to our knowledge, the first estimates of population-level behavioural responses to a future pandemic and their associations with disease risk and associated uncertainty. The inevitability of a future respiratory virus pandemic underscores the urgent need for improved pandemic preparedness and response strategies. In this context, our findings may prove useful for epidemiological forecasting and risk assessment, and to inform strategies for risk and uncertainty communication.

## Data Availability

Study data will be made available upon publication.

## Author contributions

KBP conceived of the study and acquired funding. DRMS designed the experiment and developed the survey in consultation with KBP, JB and LM. DRMS specified the model in consultation with JB and TOH. DRMS developed the code, conducted the analyses and produced the results. All authors contributed to interpretation of results. DRMS wrote the first draft. The final version of this manuscript was reviewed and approved by all authors. The underlying data were verified by DRMS, JB and KBP, and all authors had full access to the study data and accept responsibility for the decision to submit for publication.

## Data sharing

Study data will be made available upon publication.

## Acknowledgments

The authors thank all survey participants for making this work possible, as well as the six anonymous individuals who provided feedback on survey wording and clarity. DRMS and KBP are supported by the Medical Research Foundation (MRF-160-0017-ELP-POUW-C0909). The views expressed are those of the authors and not necessarily those of the institutions with which they are affiliated.

## References

1. Ahmed, F., Zviedrite, N. & Uzicanin, A. Effectiveness of workplace social distancing measures in reducing influenza transmission: a systematic review. BMC Public Health 18, 518 (2018).

2. MacIntyre, C. R. & Chughtai, A. A. Facemasks for the prevention of infection in healthcare and community settings. BMJ 350, h694 (2015).

3. Flaxman, S. et al. Estimating the effects of non-pharmaceutical interventions on COVID-19 in Europe. Nature 584, 257–261 (2020).

4. Taylor, S., Landry, C. A., Paluszek, M. M. & Asmundson, G. J. G. Reactions to COVID-19: Differential predictors of distress, avoidance, and disregard for social distancing. J. Affect. Disord. 277, 94–98 (2020).

5. Scoville, C., McCumber, A., Amironesei, R. & Jeon, J. Mask Refusal Backlash: The Politicization of Face Masks in the American Public Sphere during the Early Stages of the COVID-19 Pandemic. Socius 8, 23780231221093158 (2022).

6. Kemp, L. et al. 80 questions for UK biological security. PLOS ONE 16, e0241190 (2021).

7. Li, N. et al. General population preferences for health-related protective behaviors during infectious disease emergencies: a systematic review of conjoint-analysis studies. Soc. Sci. Med. 388, 118721 (2026).

8. Pedersen, M. J. & Favero, N. Social Distancing during the COVID-19 Pandemic: Who Are the Present and Future Noncompliers? Public Adm. Rev. 80, 805–814 (2020).

9. Taylor, S. & Asmundson, G. J. G. Negative attitudes about facemasks during the COVID-19 pandemic: The dual importance of perceived ineffectiveness and psychological reactance. PLOS ONE 16, e0246317 (2021).

10. Bavel, J. J. V. et al. Using social and behavioural science to support COVID-19 pandemic response. *Nat*. Hum. Behav. 4, 460–471 (2020).

11. Romano, A., Sotis, C., Dominioni, G. & Guidi, S. The scale of COVID-19 graphs affects understanding, attitudes, and policy preferences. Health Econ. 29, 1482–1494 (2020).

12. Louviere, J. J., Hensher, D. A. & Swait, J. D. *Stated Choice Methods: Analysis and Applications*. (Cambridge University Press, Cambridge, 2000). doi:10.1017/CBO9780511753831.

13. Cairney, P. The UK Government’s COVID-19 Policy: What Does “Guided by the Science” Mean in Practice? *Front*. Polit. Sci. 3, (2021).

14. Science Media Centre. Search results for (covid-19). https://www.sciencemediacentre.org/?s=covid-19&cat=.

15. Spiegelhalter, D. Risk and Uncertainty Communication. Annu. Rev. Stat. Its Appl. 4, 31–60 (2017).

16. Pouwels, K. B. et al. Improving the representativeness of UK’s national COVID-19 Infection Survey through spatio-temporal regression and post-stratification. Nat. Commun. 15, 5340 (2024).

17. Varia, M. et al. Investigation of a nosocomial outbreak of severe acute respiratory syndrome (SARS) in Toronto, Canada. Can. Med. Assoc. J. 169, 285–292 (2003).

18. Wong, J. Y. et al. Infection Fatality Risk of the Pandemic A(H1N1)2009 Virus in Hong Kong. Am. J. Epidemiol. 177, 834–840 (2013).

19. Eales, O. et al. Dynamics of SARS-CoV-2 infection hospitalisation and infection fatality ratios over 23 months in England. PLOS Biol. 21, e3002118 (2023).

20. Cristea, F. et al. A comparative analysis of experienced uncertainties in relation to risk communication during COVID19: a four-country study. *Glob*. Health 18, 66 (2022).

21. Wright, L., Steptoe, A. & Fancourt, D. Predictors of self-reported adherence to COVID-19 guidelines. A longitudinal observational study of 51,600 UK adults. Lancet Reg. Health – Eur. 4, (2021).

22. Dieckmann, N. F., Peters, E. & Gregory, R. At Home on the Range? Lay Interpretations of Numerical Uncertainty Ranges. Risk Anal. 35, 1281–1295 (2015).

23. Antonopoulou, V. et al. Adherence to mask wearing and social distancing and use of lateral flow tests during the COVID19 pandemic in England and Wales based on the virus watch community cohort study. *Discov*. Public Health 23, 47 (2026).

24. Binka, M., et al. The Impact of Mask Mandates on Face Mask Use During the COVID-19 Pandemic: Longitudinal Survey Study. JMIR Public Health Surveill. 9, e42616 (2023).

25. van der Bles, A. M., van der Linden, S., Freeman, A. L. J. & Spiegelhalter, D. J. The effects of communicating uncertainty on public trust in facts and numbers. Proc. Natl. Acad. Sci. 117, 7672–7683 (2020).

26. Kerr, J. et al. The effects of communicating uncertainty around statistics, on public trust. R. Soc. Open Sci. 10, 230604 (2023).

27. Smith, L. E. et al. Factors associated with adherence to self-isolation and lockdown measures in the UK: a cross-sectional survey. Public Health 187, 41–52 (2020).

28. Schneider, C. R., Freeman, A. L. J., Spiegelhalter, D. & Linden, S. van der. The effects of quality of evidence communication on perception of public health information about COVID-19: Two randomised controlled trials. PLOS ONE 16, e0259048 (2021).

29. Office for National Statistics. Average household income, UK: Financial Year Ending 2024. https://www.ons.gov.uk/peoplepopulationandcommunity/personalandhouseholdfinances/incomeandwealth/bulletins/householddisposableincomeandinequality/financialyearending2024.

30. 30. Office for National Statistics. Population of England and Wales. https://www.ethnicity-facts-figures.service.gov.uk/uk-population-by-ethnicity/national-and-regional-populations/population-of-england-and-wales/latest/?utm_source=chatgpt.com (2022).

