## Supplementary files for "Intention of UK residents to wear facemasks and practise social distancing during the next respiratory virus pandemic"

### Supplementary appendix

*Derivation of attribute levels*

Risk is commonly defined as a function of the probability of an adverse outcome and the magnitude of its consequences.[1] DCE experimental design guidelines highlight the importance of such interaction effects, for example with respect to choices to reduce pain: individuals can only make informed choices if they understand the probability of experiencing pain, its severity and its expected duration.[2] We therefore considered it critical to include both disease *prevalence* (risk of acquiring infection) and disease *severity* (risk of consequences once infected) as attributes in this DCE. Acute viral infections tend to last only from a few days to a few weeks, so infection duration was excluded, as it likely contributes less to perceived risk of a newly circulating epidemic virus than the perceived probability of getting infected and the perceived probability of poor outcomes once infected.

Both infection-hospitalisation risk (IHR) and infection-fatality risk (IFR) are key indicators of infection severity but are highly correlated. For parsimony and to avoid redundancy, a single *severity* attribute was defined that incorporates both IHR and IFR by assuming a fixed proportional relationship between them. Specifically, IFRs were informed by estimates from the literature, while IHRs were calculated relative to IFRs by assuming a 25% fatality rate among infected individuals admitted to hospital. This reflects a simplified and internally consistent relationship between hospitalisation and mortality risk for the purposes of the experiment. Examples of IFR estimates for major epidemic pathogens over recent years include 0.67% for SARS-CoV-2 in England in 2020,[3] 1.1% for influenza A(H1N1)pdm09 among older adults in Hong Kong in 2009,[4] and 13% for SARS-CoV-1 among a nosocomial cluster in Toronto, Canada in 2003.[5] Based on these values and to provide approximately proportional increases across a plausible range, severity levels (IFRs) of 0.7%, 2.0% and 6.0% were selected, with corresponding IHR values derived accordingly. See **Table S1** for the IHR and IFR values selected for low, medium and high severity levels.

For infection risk, local infection incidence and local infection prevalence are two key indicators. A single *prevalence* attribute was selected, as prevalence more directly reflects the probability that an individual's contacts are infected, and because it is generally easier to interpret than incidence rates. Levels of prevalence were informed by empirical data from the COVID-19 pandemic, including estimates of SARS-CoV-2 infection prevalence from the COVID-19 Infection Survey.[6] National prevalence in England (based on post-stratified estimates of swab positivity) reached approximately

7% over successive waves in early 2022, shortly after the emergence of the Omicron variant.[7] However, local outbreaks in small populations can reach much higher prevalence than national averages; for instance, an estimated 13% (95%CI 11%, 16%) in Tower Hamlets, London, at the start of 2022. To cover this range, values of 1.5%, 7% and 16% were selected as prevalence attribute levels.

Building on experience from the COVID-19 pandemic, it was identified that uncertainty in estimates of disease risk, and not only mean estimates, may influence behaviour,[8,9] and a range of studies have described the importance and challenges of communicating uncertainty to inform the population in a public health context.[10–12] We therefore considered *severity uncertainty* and *prevalence uncertainty* as attributes. To communicate uncertainty in risk (prevalence and severity) in the survey, it was necessary to translate categorical levels of uncertainty (low, moderate, high) into quantitative values corresponding to each included level of true risk. To make these values realistic, a simulation framework was used to represent an epidemiological study in an early pandemic context, with uncertainty quantification depending on an underlying number of samples representing realistic availability of local epidemiological data in an early epidemic context. Given binary outcomes (infected or not, hospitalised or not, deceased or not), a binomial distribution was used to quantify uncertainty from different numbers of samples (n=20, n=80, n=360). True risk was multiplied by sample size to derive a hypothetical number of positives for each outcome, and from these a 1-sample binomial proportions test was used to derive 95% confidence intervals (**Table S1**).

Following recommendations from Spiegelhalter *et al.*, severity and prevalence risks were expressed as frequencies per 1,000 population and bar charts with ranges were used to communicate risk and associated uncertainty.[1,13] In an experimental study by Dieckmann *et al.*, it was identified that the reporting of uncertainty as a numerical range can lead to different interpretations among lay people, and that visualising the range could help to improve interpretation.[14] We therefore sought to evaluate whether the inclusion of a graphical representation of prevalence and severity could impact choices, and included *text only* and *text + visualisation* as levels for the attribute *risk communication*.

***Table S1**. Derivation of levels of uncertainty in disease prevalence and severity based on the true risk level and the number of samples available from the population.*

| Risk measure | True risk level | Uncertainty level | Number of samples | Uncertainty (95% CI) |
|---|---|---|---|---|
| Prevalence (% of population with disease) | 1.5% | High | 20 | 0.1%, 18.5% |
| | | Moderate | 80 | 0.3%, 7.1% |
| | | Low | 360 | 0.7%, 3.4% |
| | 7% | High | 20 | 1.6%, 26.3% |
| | | Moderate | 80 | 3.2%, 14.8% |
| | | Low | 360 | 4.8%, 10.1% |
| | 16% | High | 20 | 5.8%, 37.2% |
| | | Moderate | 80 | 9.6%, 25.6% |
| | | Low | 360 | 12.6%, 20.1% |
| Severity (infection-hospitalisation risk) | 2.7% | High | 20 | 0.3%, 20.3% |
| | | Moderate | 80 | 0.8%, 8.9% |
| | | Low | 360 | 1.4%, 4.9% |
| | 8.0% | High | 20 | 1.9%, 27.6% |
| | | Moderate | 80 | 3.8%, 16.0% |
| | | Low | 360 | 5.6%, 11.3% |
| | 24% | High | 20 | 10.5%, 45.8% |
| | | Moderate | 80 | 16%, 34.4% |
| | | Low | 360 | 19.9%, 28.7% |
| Severity (infection-fatality risk) | 0.7% | High | 20 | <0.1%, 17.2% |
| | | Moderate | 80 | 0.1%, 5.8% |
| | | Low | 360 | 0.2%, 2.2% |
| | 2.0% | High | 20 | 0.2%, 19.3% |
| | | Moderate | 80 | 0.5%, 7.9% |
| | | Low | 360 | 1.0%, 4.0% |
| | 6.0% | High | 20 | 1.2%, 25% |
| | | Moderate | 80 | 2.5%, 13.5% |
| | | Low | 360 | 4.0%, 8.9% |

*Survey structure*

The survey was structured into seven main parts. First, participants clicking on the survey link were sent to an introductory landing page with background information and survey context, as well as ethical information and a participation consent form. Second, participants were asked to answer an initial question about their facemask wearing and social distancing practices over the month prior to the survey. Third, participants were presented with a page describing the hypothetical pandemic scenario underlying the study, including information about Disease-X transmission and symptoms, the outbreak's history, and official responses of public health organisations. Fourth, participants were presented with an explanation of the *severity* and *prevalence* of Disease-X, and were provided with examples of what these quantities mean and how they can be interpreted. Fifth, participants were shown the choices that they would be asked to make regarding facemask wearing and social distancing under different scenarios of severity and prevalence. Sixth, participants completed the DCE by making choices across each of the twelve choice tasks. Finally, participants were asked to provide information about their individual-level characteristics and were invited to provide written feedback.

The structure and wording of the survey were developed iteratively with multiple rounds of revision and editing to ensure clarity and consistency. The individual-level characteristics that participants were asked to provide were selected based on prior evidence and plausibility of potential impacts on facemask wearing and social distancing behaviours.[15,16] Recommendations from the risk communication literature were used to develop survey wording around the communication of severity and prevalence estimates, and the interpretation of uncertainty therein.[1,10,14,17] Six members of the general public with no expertise in epidemiology, statistics or health provided feedback on drafts of the survey and ensured accessibility and comprehension to a wide audience. This group included three women and three men aged 28 to 68, including one second-language and one third-language English speaker. Feedback from pilot survey participants was also used to refine the survey's wording and structure.

*Experimental design*

An initial pilot design was developed using the software Ngene (version 1.4.0). First, a multinomial logit (MNL) model was specified, incorporating an indirect utility function, including both deterministic and stochastic utility components,

$$V_{ijc} = \beta_j \cdot X'_{ic} + \varepsilon_{ijc}$$

where $V_{ijc}$ is the indirect utility of individual *i* from alternative *j* in choice task *c*; $\beta_j \cdot X'_{ic}$ is the observed utility comprising included attributes, $X'_{ic}$, and individual utility weights (or preferences) for those attributes, $\beta_j$; and $\varepsilon_{ijc}$ is the unobserved component of utility. This observed utility component was defined as,

$$\beta_j \cdot X'_{ic} = \alpha_j + \beta_{sev,j} \cdot sev_{ic} + \beta_{sevun,j} \cdot sev\ uncert_{ic} + \beta_{prev,j} \cdot prev_{ic} + \beta_{prevun,j} \cdot prev\ uncert_{ic} + \beta_{vis,j} \cdot visual_{ic}$$

where $\alpha_j$ is an alternative-specific constant (ASC) for each level of pandemic behaviour coded on a scale ranging from *j*=0 (rarely or never wear a facemask or practise social distancing) to *j*=3 (wear a facemask or practise social distancing as much as possible), where *j*=0 was taken as the reference. (See **Table 2** in the main text for corresponding levels for each attribute.) All attributes were specified as categorical and dummy coded, with the lowest category treated as the reference and omitted. Coefficients for severity ($\beta_{sev,j}$) and prevalence ($\beta_{prev,j}$) describe marginal utility associated with alternative *j* (relative to *j*=0) as a function of the severity and prevalence of Disease-X (omitting low severity and low prevalence). Coefficients for severity uncertainty ($\beta_{sevun,j}$) and prevalence uncertainty ($\beta_{prevun,j}$) describe marginal utility associated with alternative *j* given corresponding levels of uncertainty in the true severity of Disease-X and in the true prevalence of Disease-X (omitting low severity uncertainty and low prevalence uncertainty). Finally, coefficients for risk communication ($\beta_{vis,j}$) describe marginal utility associated with alternative $j$ given risk being communicated using both text and visualisation (omitting risk communication using only text).

Uninformative priors were assumed for $\alpha_j$, $\beta_{sevun,j}$, $\beta_{prevun,j}$ and $\beta_{vis,j}$, and vaguely informative priors for $\beta_{sev,j}$ and $\beta_{prev,j}$, the latter reflecting assume increased probability of mask wearing and social distancing given greater severity and prevalence. For high severity and prevalence, priors were defined to indicate a positive sign ($\beta \sim U[0.0001,1]$ for the high attribute level, $\beta \sim U[0.0001,0.5]$ for the intermediate level, and none for the low level due to dummy coding); for moderate severity and prevalence, neutrality was assumed ($\beta = 0$ for both levels); and for low severity and prevalence, a negative sign was assumed ($\beta \sim U[-1, -0.0001]$ and $\beta \sim U[-0.5, -0.0001]$ for the high and intermediate levels, respectively). A D-optimal design with 12 choice tasks and balanced levels was identified using the modified Fedorov algorithm. This pilot design was used to generate an initial pilot survey (see below for more detail on survey development). Due to adjustments to wording and design made for the final survey, pilot survey responses were not carried forward to the main analysis.

The pilot survey was launched on 08/04/2025 and 51 complete survey responses were ascertained (see below for details on survey administration). Model parameters were estimated from these 51 responses in R (version 4.4.2) using the package Apollo,[18] with separate estimation for social distancing choices and facemask wearing choices (joint estimation of simultaneous choices was only considered for the final model, see below). For more stable estimation during model fitting and to reduce the number of parameters, the variables *sev* and *prev* were considered as continuous and therefore not dummy coded. Parameter estimates for the pilot model fit to facemask wearing choices are shown in **Table S2**.

Pilot parameter estimates were used as informative Bayesian priors for estimation of the final experimental design, assuming Gaussian distributions and setting standard deviations as half means to reflect moderate uncertainty. For the final survey, the MNL model was specified with severity (*sev*) and prevalence (*prev*) treated as continuous variables, where *sev* was coded using the infection-hospitalisation ratio. A D-optimal design with 12 choice tasks and balanced levels was identified using the modified Fedorov algorithm (**Table S3**). The corresponding presentation of attribute levels is shown in **Table S4**.

***Table S2**. Multinomial logit (MNL) choice model estimates from pilot survey data (n=51) and a model specification specific to facemask wearing choices. Severity and prevalence attributes were considered as continuous variables, while severity uncertainty, prevalence uncertainty and risk communication attributes were considered as categorical variables with dummy-coding.*

| Parameter | Estimate | Std. Error | t-ratio | Robust Std. Error | Robust t-ratio |
|---|---|---|---|---|---|
| $\alpha_1$ | -0.00755 | 1.114976 | -0.00677 | 0.757882 | -0.00996 |
| $\alpha_2$ | 0.419962 | 1.118039 | 0.375624 | 0.784878 | 0.535066 |
| $\alpha_3$ | -0.00852 | 1.138452 | -0.00749 | 0.766444 | -0.01112 |
| $\beta_{sev,1}$ | 0.009049 | 0.004577 | 1.976949 | 0.00534 | 1.694428 |
| $\beta_{sev,2}$ | 0.009773 | 0.0045 | 2.171981 | 0.005666 | 1.724775 |
| $\beta_{sev,3}$ | 0.011237 | 0.004485 | 2.505467 | 0.005351 | 2.099909 |
| $\beta_{prev,1}$ | 0.008512 | 0.006106 | 1.393995 | 0.005379 | 1.582279 |
| $\beta_{prev,2}$ | 0.009186 | 0.005989 | 1.533819 | 0.005795 | 1.585058 |
| $\beta_{prev,3}$ | 0.011988 | 0.00596 | 2.011201 | 0.005482 | 2.186578 |
| $\beta_{modsevun,1}$ | -0.19003 | 0.91451 | -0.20779 | 0.814339 | -0.23335 |
| $\beta_{highsevun,1}$ | -0.54613 | 0.951888 | -0.57373 | 0.747613 | -0.7305 |
| $\beta_{modsevun,2}$ | -0.27376 | 0.894348 | -0.3061 | 0.770194 | -0.35544 |
| $\beta_{highsevun,2}$ | -0.55506 | 0.930671 | -0.5964 | 0.649781 | -0.85422 |
| $\beta_{modsevun,3}$ | 0.257507 | 0.8904 | 0.289204 | 0.746612 | 0.344901 |
| $\beta_{highsevun,3}$ | -0.14161 | 0.927922 | -0.15261 | 0.654562 | -0.21634 |
| $\beta_{modprevun,1}$ | 0.455614 | 0.931277 | 0.489235 | 1.114407 | 0.408839 |
| $\beta_{highprevun,1}$ | 0.799976 | 0.742581 | 1.077291 | 0.321501 | 2.488251 |
| $\beta_{modprevun,2}$ | 0.295495 | 0.915051 | 0.322927 | 1.065452 | 0.277342 |
| $\beta_{highprevun,2}$ | 0.870484 | 0.720948 | 1.207416 | 0.294951 | 2.951288 |
| $\beta_{modprevun,3}$ | 0.498533 | 0.911819 | 0.546745 | 1.059125 | 0.470702 |
| $\beta_{highprevun,3}$ | 0.76947 | 0.715107 | 1.07602 | 0.274958 | 2.7985 |
| $\beta_{vis,1}$ | 0.333834 | 0.648635 | 0.514672 | 0.531124 | 0.628543 |
| $\beta_{vis,2}$ | 0.468585 | 0.632188 | 0.741211 | 0.438658 | 1.068224 |
| $\beta_{vis,3}$ | 0.153051 | 0.627467 | 0.243919 | 0.434317 | 0.352395 |

***Table S3***. *Final survey design.*

| Task | **Severity** (hospitalisations/ 1,000 infections) | **Severity uncertainty** | **Prevalence** (infections/ 1,000 population) | **Prevalence uncertainty** | **Risk communication** |
|---|---|---|---|---|---|
| 1 | High (240) | High | Low (15) | Low | Only text |
| 2 | Low (27) | Low | Low (15) | Moderate | Only text |
| 3 | Low (27) | High | High (160) | Low | Only text |
| 4 | Moderate (80) | Moderate | High (160) | Moderate | Only text |
| 5 | High (240) | Low | Moderate (70) | High | Only text |
| 6 | Moderate (80) | Moderate | Moderate (70) | High | Only text |
| 7 | Low (27) | Moderate | Low (15) | Low | Text + visual |
| 8 | High (240) | Moderate | Moderate (70) | Moderate | Text + visual |
| 9 | Low (27) | High | Low (15) | High | Text + visual |
| 10 | High (240) | Low | High (160) | High | Text + visual |
| 11 | Moderate (80) | High | Moderate (70) | Moderate | Text + visual |
| 12 | Moderate (80) | Low | High (160) | Low | Text + visual |

***Table S4***. *Final survey design as presented to survey participants. Uncertainty intervals are provided in parentheses. Detailed information on interpretation of these numbers was provided to survey participants (see full survey text online).*

| Task | Infections / 1,000 population | Hospitalisations / 1,000 infections | Deaths / 1,000 infections | Risk communication |
|---|---|---|---|---|
| 1 | 15 (7, 34) | 240 (105, 458) | 60 (12, 250) | Only text |
| 2 | 15 (3, 71) | 27 (14, 49) | 7 (2, 22) | Only text |
| 3 | 160 (126, 201) | 27 (3, 203) | 7 (<1, 172) | Only text |
| 4 | 160 (96, 256) | 80 (38, 160) | 20 (5, 79) | Only text |
| 5 | 70 (16, 263) | 240 (199, 287) | 60 (40, 89) | Only text |
| 6 | 70 (16, 263) | 80 (38, 160) | 20 (5, 79) | Only text |
| 7 | 15 (7, 34) | 27 (8, 89) | 7 (1, 58) | Text + visual |
| 8 | 70 (32, 148) | 240 (160, 344) | 60 (25, 135) | Text + visual |
| 9 | 15 (1, 185) | 27 (3, 203) | 7 (<1, 172) | Text + visual |
| 10 | 160 (58, 372) | 240 (199, 287) | 60 (40, 89) | Text + visual |
| 11 | 70 (32, 148) | 80 (19, 276) | 20 (2, 193) | Text + visual |
| 12 | 160 (126, 201) | 80 (56, 113) | 20 (10, 40) | Text + visual |

*Survey administration*

Three survey components were randomised at the individual participant level. First, the survey was split into two blocks by the *risk communication* attribute with block order randomised, such that participants were shown six consecutive tasks with one attribute level (e.g. text only) followed by six consecutive tasks with the other (e.g. text + visual). The decision to block the presentation variable was made after concern arose during survey testing that some participants interpreted a randomized appearance of images as a technical glitch. Second, within each of these blocks, the order of choice tasks was randomized. Third, within each choice task the order of appearance of the two principal disease characteristics (severity and prevalence) was randomized, as was the order of the two behaviours to choose from (mask wearing and social distancing).

***Table S5***. *Survey quotas. Note that quotas contain initial full sample (n=2,008) before the removal of two respondents with data quality issues.*

| | 2021 Census (%) | Target quota (N) | Respondents (N) |
|---|---|---|---|
| ***Gender*** | | | |
| Male | 48.3 | 966 | 968 |
| Female | 51.7 | 1,034 | 1,034 |
| Other / Prefer not to say | --- | 0 | 6 |
| **Total** | **100** | **2,000** | **2,008** |
| ***Age*** | | | |
| 18-24 | 10.7 | 214 | 216 |
| 25-34 | 16.9 | 338 | 340 |
| 35-44 | 16.4 | 328 | 330 |
| 45-54 | 16.9 | 338 | 337 |
| 55-64 | 15.9 | 318 | 319 |
| 65+ | 23.2 | 464 | 466 |
| **Total** | **100** | **2,000** | **2,008** |
| ***Region*** | | | |
| East Midlands | 7.4 | 148 | 150 |
| East of England | 9.0 | 180 | 181 |
| Greater London | 11.9 | 238 | 239 |
| North East England | 4.1 | 82 | 82 |
| North West England | 10.9 | 218 | 219 |
| Northern Ireland | 2.8 | 56 | 56 |
| Scotland | 8.0 | 160 | 161 |
| South East England | 15.9 | 318 | 319 |
| South West England | 8.2 | 164 | 164 |
| Wales | 4.7 | 94 | 95 |
| West Midlands | 8.7 | 174 | 174 |
| Yorkshire and the Humber | 8.4 | 168 | 168 |
| **Total** | **100** | **2,000** | **2,008** |

*Survey responses*

After consenting to participate in the survey, and before being provided with survey context describing the hypothetical pandemic scenario under consideration, participants were asked to describe their mask wearing and social distancing practises over the month prior to the survey (**Figure S2**). All pairwise survey responses for each choice task in the DCE (pairwise mask wearing choices and social distancing choices) are provided in **Figure S1**.

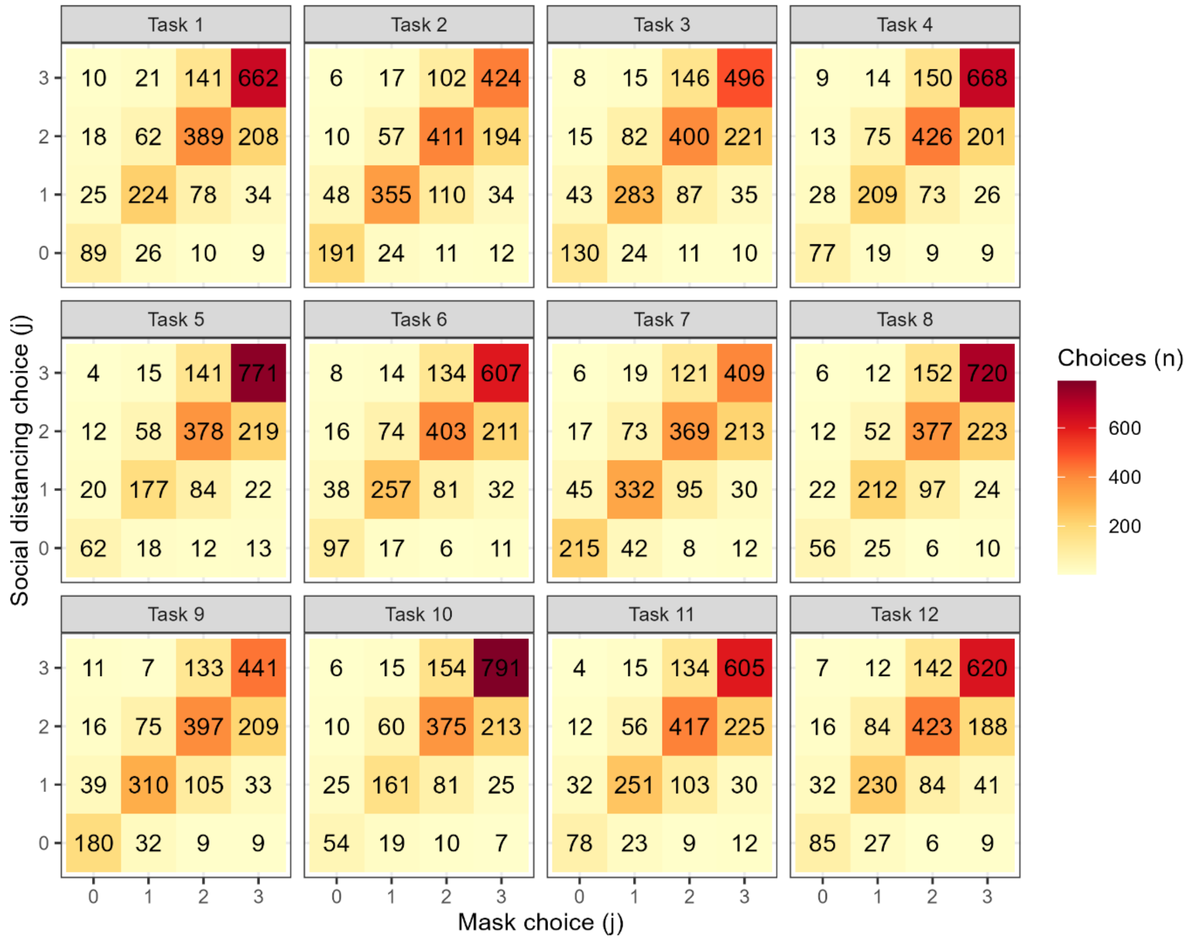


***Figure S1***. *Pairwise choices for mask wearing (x-axis) and social distancing (y-axis) in each choice task. Choices in each task sum to 2,006 unique responses.*

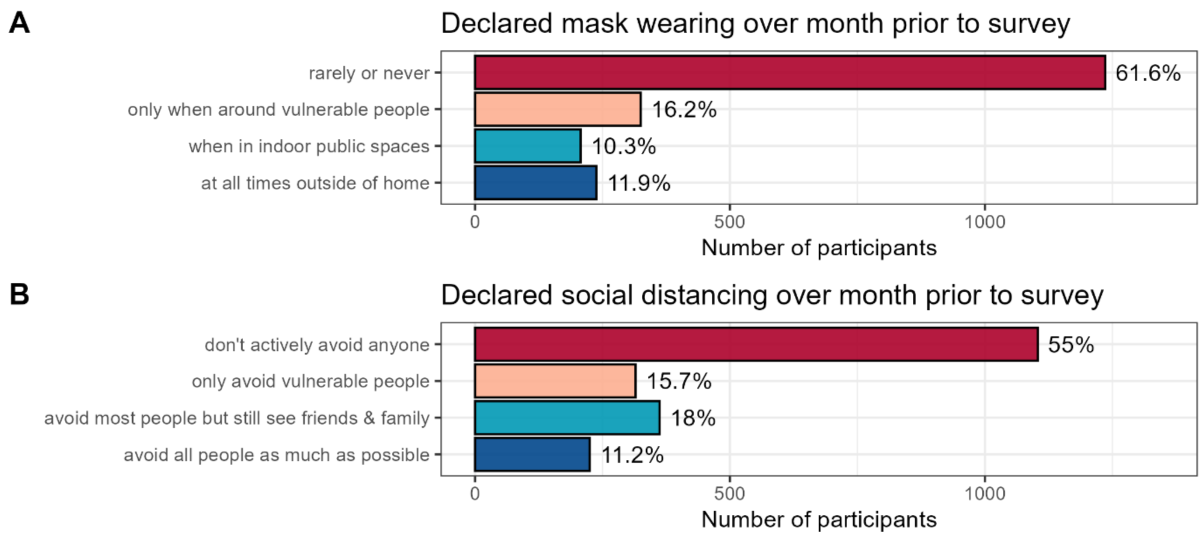


***Figure S2***. *Declared current behaviours over the month prior to participating in the survey among survey participants (n=2,006).*

*Covariates*

After completing the DCE, participants provided information about their individual-level sociodemographic characteristics (covariates). Most of these questions included multiple categorical response options. However, including each category level as a separate dummy variable led to unstable model estimation and, in some instances, model non-convergence, in particular due to small numbers of participants responding to certain categories like "prefer not to say" (PNTS). [On average, 22.1 (1.1%) participants responded PNTS across the 10 questions including PNTS as an option.] To avoid over-parameterisation and unstable estimation due to sparse categories, it was necessary to collapse levels of categorical variables where appropriate to ensure sufficient observations within each category (**Table S6**). The covariate *number of household members* was excluded due to data quality concerns: upon reviewing participant comments, 4 independently stated that they may have misunderstood this question and were unsure if their answer was correct.

The average number of individuals choosing PNTS was low for each question, but, overall, 8.4% (n=168) of participants responded PNTS to at least one question. A sensitivity analysis was therefore conducted, removing all 168 individuals who answered PNTS to any question, and re-estimating the model among the remaining 91.6% of the survey sample. Such a complete case analysis is unbiased under the assumption that the missingness in PNTS responses is not jointly dependent on both the chosen alternatives and the attribute levels.[19]

***Table S6***. *Collapsed levels of sociodemographic covariates considered in model specification. See* ***Table 3*** *for levels included in "all other" for each covariate.*

| Covariate | Collapsed levels |
|---|---|
| Age | <40 years<br>40-59 years<br>60+ years |
| Gender | Female<br>All other |
| Region | England<br>Scotland, Northern Ireland or Wales |
| Current behaviour | Not cautious ($j = 0$)<br>Cautious ($j \in \{3,2,1\}$) |
| Area of residence | City (urban)<br>City (suburban)<br>Town<br>All other |
| Highest education | University degree<br>All other |
| Number of times vaccinated against COVID-19 | 4 or more times<br>1 to 3 times<br>All other |
| Health or social care worker | Yes or used to be<br>All other |
| Gross annual household income | £75,000 or more<br>£40,000 to £74,999<br>£20,000 to £39,999<br>All other |
| Living with serious chronic illness | Yes<br>All other |
| Living with or caring for fragile or vulnerable people | Yes<br>All other |
| Days per week working from home | 1 or more<br>All other |
| Ethnicity | White<br>All other |

*Model specification*

For each behaviour *b*, the deterministic component of utility specifies the systematic utility of alternative *j* for individual *i* in choice task *c*. It includes $\alpha_j^{(b)}$, the alternative-specific constant (ASC) term, and expands to

$$\beta_j^{(b)} \cdot X'_{ic} = \alpha_j^{(b)} + \beta_{sev,j}^{(b)} \cdot sev_{ic} + \beta_{modsevun,j}^{(b)} \cdot mod\ sev\ uncert_{ic} + \beta_{highsevun,j}^{(b)} \cdot high\ sev\ uncert_{ic} + \beta_{prev,j}^{(b)} \cdot prev_{ic} + \beta_{modprevun,j}^{(b)} \cdot mod\ prev\ uncert_{ic} + \beta_{highprevun,j}^{(b)} \cdot high\ prev\ uncert_{ic} + \beta_{vis,j}^{(b)} \cdot visual_{ic}$$

The ASC term is composed of an intercept term $\beta_{int}$ and a linear combination of individual-level covariates, the product of a vector of coefficients on the covariates and a vector of the covariates themselves (see below for more details on covariate selection). Note that *current behaviour* is the only covariate specified as behaviour-specific: it was assumed that facemask wearing behaviour in the month prior to the survey had an impact only on facemask wearing choices, and that social distancing behaviour over the month prior to the survey had an impact only on social distancing choices.

The severity attribute is expressed on a continuous scale of $sev$ hospitalisations per 1,000 Disease-X infections, and the prevalence attribute is expressed on a continuous scale of $prev$ Disease-X infections per 1,000 population. The severity uncertainty, prevalence uncertainty and risk communication attributes are dummy-coded, each having one level omitted to allow for model identification (respectively: low severity uncertainty, low prevalence uncertainty, and risk communication using only text). To identify the model, the base alternative describing least masking or distancing (*j*=0) was normalised such that all coefficients for categorical variables are interpreted relative to the base alternative:

$$\alpha_0^{(b)} = \beta_{modsevun,0}^{(b)} = \beta_{highsevun,0}^{(b)} = \beta_{modprevun,0}^{(b)} = \beta_{highprevun,0}^{(b)} = \beta_{vis,0}^{(b)} = 0$$

For individual *i* choosing alternative *j* in choice task *c*, conditional on their vector of latent affinities, the probability of choice *j* takes the standard MNL form,

$$P\left(y_{i,c}^{(b)} = j \mid \eta_i\right) = \frac{\exp\left(\beta_j^{(b)} \cdot X'_{ic} + \eta_{i,j}\right)}{\sum_{k=0}^{3} \exp\left(\beta_k^{(b)} \cdot X'_{ic} + \eta_{i,j}\right)}$$

where $\eta_{i,0} = 0$ for the base alternative (normalisation).

### *Model selection*

To account for residual preference heterogeneity beyond included attributes, intercepts and random error terms, data collected on individual-level covariates were considered for inclusion in model specification. To avoid an over-parameterised model while identifying covariates that improve model fit, an iterative forwards-backwards model specification process was conducted to evaluate which combinations of covariates best improve overall model fit (**Table S7**). Starting with a baseline model that included only attributes and intercept terms, covariates from the list were added sequentially in a forward-stepwise manner, and the tested covariate was carried forward only if the resulting BIC was lower than the previous model. Then, once all covariates were tested, each covariate was sequentially removed in a backwards-stepwise manner, to test whether an included covariate no longer improved model fit after other covariates had been added. Finally, after this forwards-backwards procedure was complete, selected covariates were tested for linear interactions with severity and prevalence.

Covariates are interpreted as follows, relative to terms omitted due to dummy coding: aged 40 to 59 years ($midage$) or aged 60+ years ($oldage$) relative to aged <40 years (omitted); any amount of facemask wearing or social distancing over the month prior to the survey ($basebehav$) relative to none (omitted); a university degree or equivalent ($education$) relative to less educational attainment (omitted); previous receipt of 1 to 3 COVID-19 vaccinations ($vaxfew$) or 4+ vaccinations ($vaxmany$) relative to none (omitted); current or previous experience as a health and social care worker ($hscw$) relative to none (omitted); suffering from a serious chronic illness ($chronic$) relative to not (omitted); living with or caring for any fragile or vulnerable people ($caretaking$) relative to not (omitted); and white ethnicity ($white$) relative to other ethnicity (omitted).

***Table S7***. *Model selection procedure for the primary analysis. Model 20 (in bold) resulted in the lowest BIC and was selected. X indicates inclusion of a covariate in the model. S indicates inclusion of a covariate and its linear interaction with severity. P indicates inclusion of a covariate and its linear interaction with prevalence. B indicates inclusion of a covariate and its linear interactions with both severity and prevalence. HSCW = health and social care worker; WFH = working from home; LL = log-likelihood; BIC = Bayesian information criterion.*

| | Covariates included | | | | | | | | | | | | | | |
|---|---|---|---|---|---|---|---|---|---|---|---|---|---|---|---|
| **Model** | *Age* | *Sex* | *Current behaviour* | *Area of residence* | *Region* | *Highest education* | *COVID-19 vaccination* | *HSCW* | *Household income* | *Chronic illness* | *Vulnerable contacts* | *Days WFH* | *Ethnicity* | **LL** | **BIC** |
| 1 | | | | | | | | | | | | | | -37725.64 | 75965.81 |
| 2 | X | | | | | | | | | | | | | -37555.55 | 75746.7 |
| 3 | X | X | | | | | | | | | | | | -37549.2 | 75794.52 |
| 4 | X | | X | | | | | | | | | | | -37300.62 | 75297.37 |
| 5 | X | | X | X | | | | | | | | | | -37235.32 | 75348.37 |
| 6 | X | | X | | X | | | | | | | | | -37295.41 | 75347.48 |
| 7 | X | | X | | | X | | | | | | | | -37243.27 | 75243.2 |
| 8 | X | | X | | | X | X | | | | | | | -37104.26 | 75086.24 |
| 9 | X | | X | | | X | X | X | | | | | | -37094.9 | 75128.07 |
| 10 | X | | X | | | X | X | | X | | | | | -37092.88 | 75124.01 |
| 11 | X | | X | | | X | X | | | X | | | | -37083.33 | 75104.92 |
| 12 | X | | X | | | X | X | | | | X | | | -37089.25 | 75116.75 |
| 13 | X | | X | | | X | X | | | | | X | | -37095.55 | 75129.36 |
| 14 | X | | X | | | X | X | | | | | | X | -37052.41 | 75043.08 |
| 15 | | | X | | | X | X | | | | | | X | -37109.9 | 75036.98 |
| 16 | | | | | | X | X | | | | | | X | -37353.26 | 75463.18 |
| 17 | | | X | | | | X | | | | | | X | -37159.2 | 75075.05 |
| 18 | | | X | | | X | | | | | | | X | -37273.7 | 75243.53 |
| 19 | | | X | | | X | X | | | | | | | -37154.32 | 75065.31 |
| **20** | | | **S** | | | **X** | **X** | | | | | | **X** | **-37034.72** | **74947.16** |
| 21 | | | B | | | X | X | | | | | | X | -37027.46 | 74993.19 |
| 22 | | | S | | | S | X | | | | | | X | -37029.26 | 74996.78 |
| 23 | | | S | | | P | X | | | | | | X | -37034.36 | 75006.98 |
| 24 | | | S | | | X | S | | | | | | X | -37002.8 | 75004.39 |
| 25 | | | S | | | X | P | | | | | | X | -37015.31 | 75029.4 |
| 26 | | | S | | | X | X | | | | | | S | -37006.11 | 74950.48 |
| 27 | | | S | | | X | X | | | | | | P | -37018.19 | 74974.63 |

***Table S8***. *Model selection procedure for the sensitivity analysis excluding current behaviour as a covariate. Model 25 (in bold) resulted in the lowest BIC and was selected. X indicates inclusion of a covariate in the model. S indicates inclusion of a covariate and its linear interaction with severity. P indicates inclusion of a covariate and its linear interaction with prevalence. B indicates inclusion of a covariate and its linear interactions with both severity and prevalence. HSCW = health and social care worker; WFH = working from home; LL = log-likelihood; BIC = Bayesian information criterion.*

| | **Covariates included** | | | | | | | | | | | | | | |
|---|---|---|---|---|---|---|---|---|---|---|---|---|---|---|---|
| **Model** | *Age* | *Sex* | *~~Current behaviour~~* | *Area of residence* | *Region* | *Highest education* | *COVID-19 vaccination* | *HSCW* | *Household income* | *Chronic illness* | *Vulnerable contacts* | *Days WFH* | *Ethnicity* | **LL** | **BIC** |
| 1 | | | | | | | | | | | | | | -37725.64 | 75965.81 |
| 2 | X | | | | | | | | | | | | | -37555.55 | 75746.7 |
| 3 | X | X | | | | | | | | | | | | -37549.2 | 75794.52 |
| 4 | X | | | X | | | | | | | | | | -37457.97 | 75733.12 |
| 5 | X | | | X | X | | | | | | | | | -37456.1 | 75789.93 |
| 6 | X | | | X | | X | | | | | | | | -37420.97 | 75719.66 |
| 7 | X | | | X | | X | X | | | | | | | -37259.38 | 75517.55 |
| 8 | X | | | X | | X | X | X | | | | | | -37246.88 | 75553.09 |
| 9 | X | | | X | | X | X | | X | | | | | -37248.06 | 75555.45 |
| 10 | X | | | X | | X | X | | | X | | | | -37256.34 | 75572 |
| 11 | X | | | X | | X | X | | | | X | | | -37249.33 | 75557.99 |
| 12 | X | | | X | | X | X | | | | | X | | -37243.75 | 75546.81 |
| 13 | X | | | X | | X | X | | | | | | X | -37211.21 | 75481.74 |
| 14 | | | | X | | X | X | | | | | | X | -37279.13 | 75496.52 |
| 15 | X | | | | | X | X | | | | | | X | -37270.89 | 75419.51 |
| 16 | X | | | | | | X | | | | | | X | -37319.04 | 75455.28 |
| 17 | X | | | | | X | | | | | | | X | -37429.69 | 75616.04 |
| 18 | X | | | | | X | X | | | | | | | -37343.37 | 75503.94 |
| 19 | S | | | | | X | X | | | | | | | -37210.66 | 75420.11 |
| 20 | P | | | | | X | X | | | | | | X | -37251.73 | 75502.25 |
| 21 | X | | | | | S | X | | | | | | X | -37261.68 | 75461.62 |
| 22 | X | | | | | P | X | | | | | | X | -37270.4 | 75479.06 |
| 23 | X | | | | | X | S | | | | | | X | -37229.21 | 75457.22 |
| 24 | X | | | | | X | P | | | | | | X | -37251.1 | 75501 |
| **25** | **X** | | | | | **X** | **X** | | | | | | **S** | **-37215.42** | **75369.1** |
| 26 | X | | | | | X | X | | | | | | B | -37200 | 75398.8 |

***Table S9***. *Coefficients for attribute levels estimated from the main model.*

| Attribute (level) | Choice alternative | Mask wearing ($b$ = M) | | | Social distancing ($b$ = D) | | |
|---|---|---|---|---|---|---|---|
| | | Estimate (95% CI) | Robust St. Error | Robust t-ratio | Estimate (95% CI) | Robust St. Error | Robus t t-ratio |
| $\beta_{s,j}^{(b)}$ Severity (continuous) | $j$ = 0 | Reference | / | / | Reference | / | / |
| | $j$ = 1 | 0.00405 (0.00291, 0.00520) | 0.000583 | 6.96 | 0.00343 (0.00224, 0.00462) | 0.000609 | 5.63 |
| | $j$ = 2 | 0.00944 (0.00820, 0.01069) | 0.000634 | 14.89 | 0.00898 (0.00760, 0.01035) | 0.000701 | 12.80 |
| | $j$ = 3 | 0.01473 (0.01334, 0.01611) | 0.000705 | 20.88 | 0.01517 (0.01360, 0.01675) | 0.000803 | 18.89 |
| $\beta_{p,j}^{(b)}$ Prevalence (continuous) | $j$ = 0 | Reference | / | / | Reference | / | / |
| | $j$ = 1 | 0.00399 (0.00287, 0.00512) | 0.000575 | 6.95 | 0.00432 (0.00314, 0.00549) | 0.000601 | 7.18 |
| | $j$ = 2 | 0.00836 (0.00723, 0.00948) | 0.000573 | 14.58 | 0.00919 (0.00799, 0.01039) | 0.000611 | 15.04 |
| | $j$ = 3 | 0.01119 (0.00997, 0.01241) | 0.000621 | 18.02 | 0.01294 (0.01162, 0.01426) | 0.000671 | 19.27 |
| $\beta_{msu,j}^{(b)}$ Severity uncertainty (moderate) | $j$ = 0 | Reference | / | / | Reference | / | / |
| | $j$ = 1 | 0.101 (-0.042, 0.244) | 0.0729 | 1.38 | 0.095 (-0.050, 0.239) | 0.0738 | 1.28 |
| | $j$ = 2 | 0.133 (-0.003, 0.268) | 0.0690 | 1.92 | 0.264 (0.123, 0.405) | 0.0719 | 3.68 |
| | $j$ = 3 | 0.235 (0.090, 0.379) | 0.0737 | 3.19 | 0.381 (0.227, 0.534) | 0.0783 | 4.87 |
| $\beta_{hsu,j}^{(b)}$ Severity uncertainty (high) | $j$ = 0 | Reference | / | / | Reference | / | / |
| | $j$ = 1 | 0.117 (-0.025, 0.260) | 0.0728 | 1.61 | 0.151 (0.004, 0.297) | 0.0747 | 2.02 |
| | $j$ = 2 | 0.235 (0.097, 0.373) | 0.0703 | 3.34 | 0.309 (0.163, 0.454) | 0.0742 | 4.16 |
| | $j$ = 3 | 0.299 (0.150, 0.448) | 0.0760 | 3.94 | 0.393 (0.232, 0.554) | 0.0823 | 4.77 |
| $\beta_{mpu,j}^{(b)}$ Prevalence uncertainty (moderate) | $j$ = 0 | Reference | / | / | Reference | / | / |
| | $j$ = 1 | 0.337 (0.194, 0.479) | 0.0726 | 4.64 | 0.402 (0.253, 0.552) | 0.0762 | 5.28 |
| | $j$ = 2 | 0.662 (0.529, 0.795) | 0.0678 | 9.77 | 0.53 (0.38, 0.679) | 0.0762 | 6.95 |
| | $j$ = 3 | 0.781 (0.641, 0.921) | 0.0714 | 10.95 | 0.676 (0.516, 0.837) | 0.0821 | 8.24 |
| $\beta_{hpu,j}^{(b)}$ Prevalence uncertainty (high) | $j$ = 0 | Reference | / | / | Reference | / | / |
| | $j$ = 1 | 0.144 (0.000, 0.287) | 0.0733 | 1.96 | 0.227 (0.080, 0.374) | 0.0750 | 3.03 |
| | $j$ = 2 | 0.447 (0.311, 0.584) | 0.0698 | 6.41 | 0.450 (0.302, 0.599) | 0.0760 | 5.93 |
| | $j$ = 3 | 0.627 (0.484, 0.770) | 0.0730 | 8.59 | 0.635 (0.475, 0.795) | 0.0818 | 7.76 |
| $\beta_{v,j}^{(b)}$ Risk communication (text + visual) | $j$ = 0 | Reference | / | / | Reference | / | / |
| | $j$ = 1 | 0.082 (-0.054, 0.218) | 0.0692 | 1.19 | 0.012 (-0.128, 0.152) | 0.0714 | 0.17 |
| | $j$ = 2 | 0.108 (-0.029, 0.244) | 0.0696 | 1.54 | -0.023 (-0.164, 0.119) | 0.0723 | -0.31 |
| | $j$ = 3 | 0.077 (-0.072, 0.225) | 0.0758 | 1.02 | -0.059 (-0.219, 0.101) | 0.0814 | -0.72 |

***Table S10***. *Coefficients for individual-level characteristics (deterministic heterogeneity) estimated from the main model.*

| Covariate | Choice alternative | Mask wearing ($b$ = M) | | | Social distancing ($b$ = D) | | |
|---|---|---|---|---|---|---|---|
| | | Estimate (95% CI) | Robust St. Error | Robust t-ratio | Estimate (95% CI) | Robust St. Error | Robust t-ratio |
| $\beta^{(b)}_{int,j}$ Intercept | $j$ = 0 | Reference | / | / | Reference | / | / |
| | $j$ = 1 | -1.459 (-2.326, -0.591) | 0.4426 | -3.30 | -1.165 (-2.027, -0.303) | 0.4399 | -2.65 |
| | $j$ = 2 | -2.390 (-3.455, -1.325) | 0.5433 | -4.40 | -1.707 (-2.709, -0.706) | 0.5110 | -3.34 |
| | $j$ = 3 | -2.585 (-3.641, -1.529) | 0.5389 | -4.80 | -3.385 (-4.436, -2.334) | 0.5362 | -6.31 |
| $\beta^{(b)}_{basebehav,j}$ Cautious current behaviour | $j$ = 0 | Reference | / | / | Reference | / | / |
| | $j$ = 1 | 1.597 (1.058, 2.136) | 0.2748 | 5.81 | 1.391 (0.882, 1.9) | 0.2598 | 5.35 |
| | $j$ = 2 | 1.888 (1.325, 2.45) | 0.2869 | 6.58 | 1.793 (1.286, 2.299) | 0.2584 | 6.94 |
| | $j$ = 3 | 2.856 (2.261, 3.452) | 0.3039 | 9.40 | 2.464 (1.897, 3.031) | 0.2894 | 8.51 |
| $\beta^{(b)}_{education,j}$ Higher education | $j$ = 0 | Reference | / | / | Reference | / | / |
| | $j$ = 1 | 0.185 (-0.36, 0.731) | 0.2784 | 0.67 | 0.568 (0.025, 1.112) | 0.2772 | 2.05 |
| | $j$ = 2 | 0.241 (-0.268, 0.75) | 0.2598 | 0.93 | 0.388 (-0.088, 0.864) | 0.2429 | 1.60 |
| | $j$ = 3 | 0.679 (0.048, 1.309) | 0.3217 | 2.11 | 0.873 (0.271, 1.476) | 0.3074 | 2.84 |
| $\beta^{(b)}_{vaxfew,j}$ 1 to 3 COVID-19 vaccine doses | $j$ = 0 | Reference | / | / | Reference | / | / |
| | $j$ = 1 | 1.24 (0.623, 1.856) | 0.3147 | 3.94 | 0.758 (0.157, 1.359) | 0.3066 | 2.47 |
| | $j$ = 2 | 2.323 (1.496, 3.149) | 0.4217 | 5.51 | 1.508 (0.717, 2.299) | 0.4035 | 3.74 |
| | $j$ = 3 | 1.632 (0.848, 2.416) | 0.4000 | 4.08 | 1.29 (0.495, 2.084) | 0.4055 | 3.18 |
| $\beta^{(b)}_{vaxmany,j}$ 4+ COVID-19 vaccine doses | $j$ = 0 | Reference | / | / | Reference | / | / |
| | $j$ = 1 | 1.331 (0.587, 2.074) | 0.3793 | 3.51 | 0.557 (-0.153, 1.268) | 0.3625 | 1.54 |
| | $j$ = 2 | 3.087 (2.233, 3.941) | 0.4358 | 7.08 | 2.043 (1.241, 2.846) | 0.4095 | 4.99 |
| | $j$ = 3 | 3.085 (2.278, 3.892) | 0.4116 | 7.50 | 2.54 (1.751, 3.329) | 0.4026 | 6.31 |
| $\beta^{(b)}_{white,j}$ White ethnicity | $j$ = 0 | Reference | / | / | Reference | / | / |
| | $j$ = 1 | -0.299 (-0.98, 0.382) | 0.3476 | -0.86 | 0.066 (-0.584, 0.715) | 0.3313 | 0.20 |
| | $j$ = 2 | -0.162 (-0.842, 0.518) | 0.3467 | -0.47 | 0.007 (-0.623, 0.638) | 0.3217 | 0.02 |
| | $j$ = 3 | -1.729 (-2.513, -0.946) | 0.3999 | -4.32 | -0.891 (-1.656, -0.126) | 0.3904 | -2.28 |
| $\gamma^{(b)}_{sev,basebehav}$ Current behaviour and severity interaction | $j$ = 0 | Reference | / | / | Reference | / | / |
| | $j$ = 1 | -0.00138 (-0.00371, 0.00094) | 0.001186 | -1.16 | -0.0019 (-0.00416, 0.00037) | 0.001155 | -1.64 |
| | $j$ = 2 | -0.00394 (-0.00639, -0.00149) | 0.001249 | -3.15 | -0.00457 (-0.0069, -0.00224) | 0.001187 | -3.85 |
| | $j$ = 3 | -0.00642 (-0.00895, -0.00389) | 0.001291 | -4.97 | -0.00722 (-0.00972, -0.00472) | 0.001275 | -5.66 |

***Table S11***. *Random effects estimated from the main model.*

| Covariate | Choice alternative | Estimate (95% CI) | Robust St. Error | Robust t-ratio |
|---|---|---|---|---|
| $\eta_j$ Random effects | $j$ = 0 | Reference | / | / |
| | $j$ = 1 | -2.45 (-2.748, -2.152) | 0.1520 | -16.11 |
| | $j$ = 2 | 3.134 (2.839, 3.429) | 0.1504 | 20.83 |
| | $j$ = 3 | 4.832 (4.531, 5.133) | 0.1537 | 31.45 |

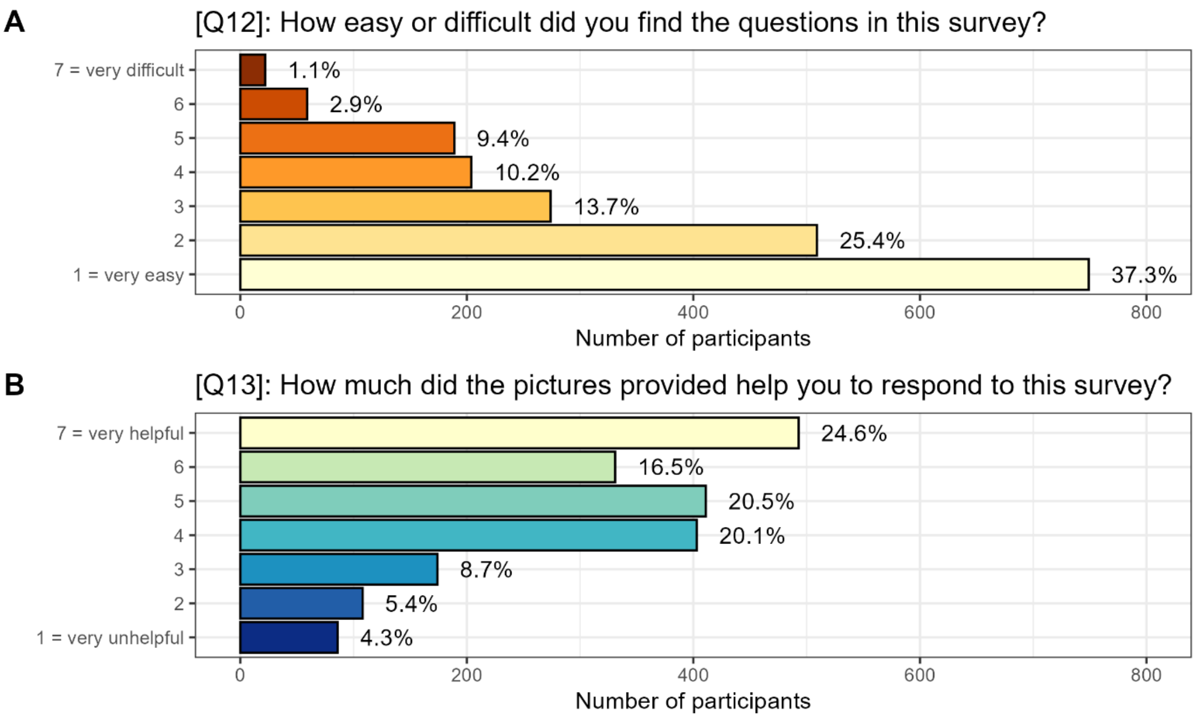


***Figure S3***. *Survey participant responses to questions regarding ease of understanding and helpfulness of the pictures provided. Both questions were answered on a Likert scale from 1 to 7.*

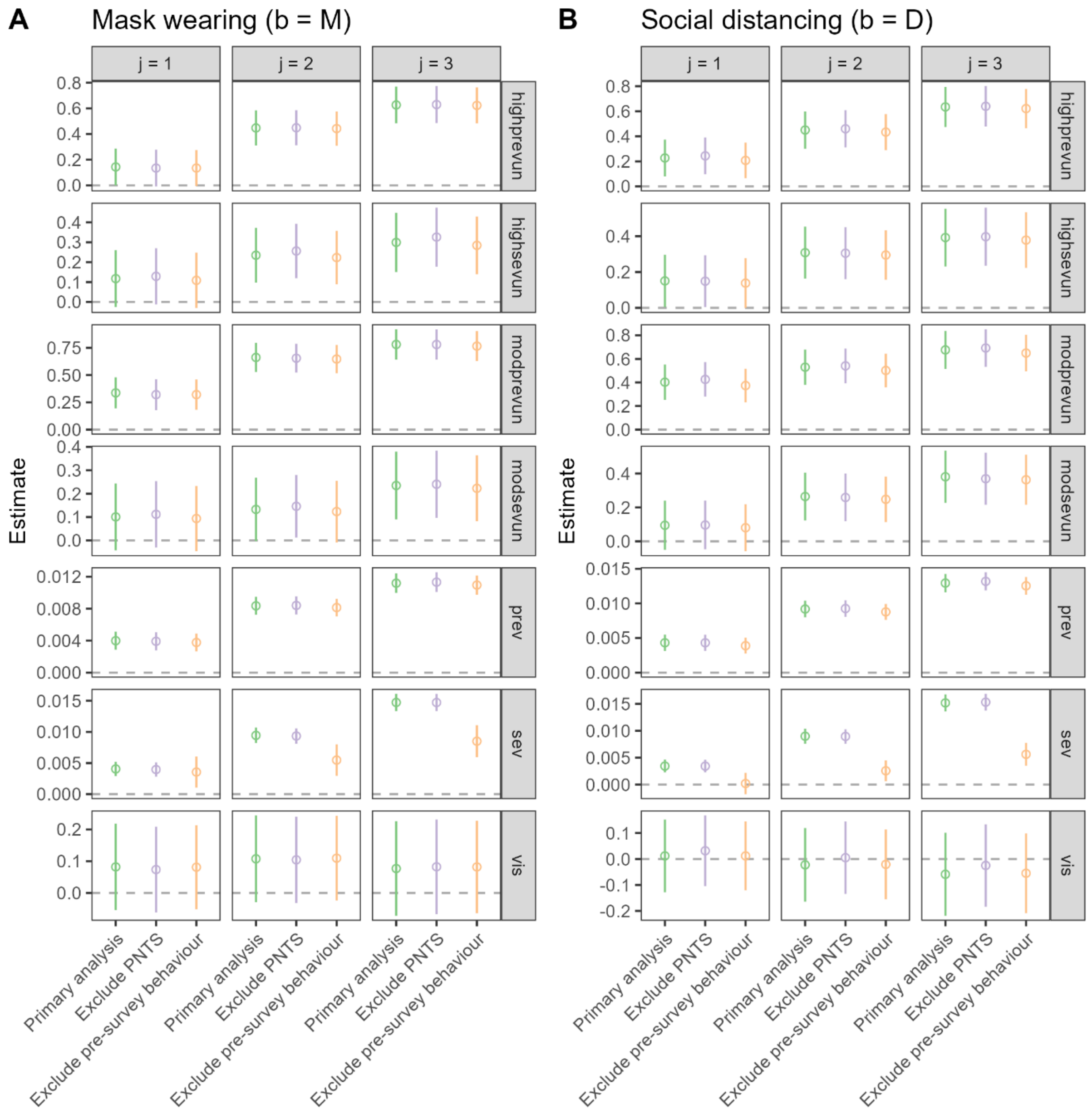


***Figure S4***. *Estimated model coefficients for model attributes, comparing the primary analysis (green) and sensitivity analysis excluding individuals responding prefer not to say (PNTS). Points and error bars represent, respectively, means and 95% confidence intervals estimated from robust standard errors.*

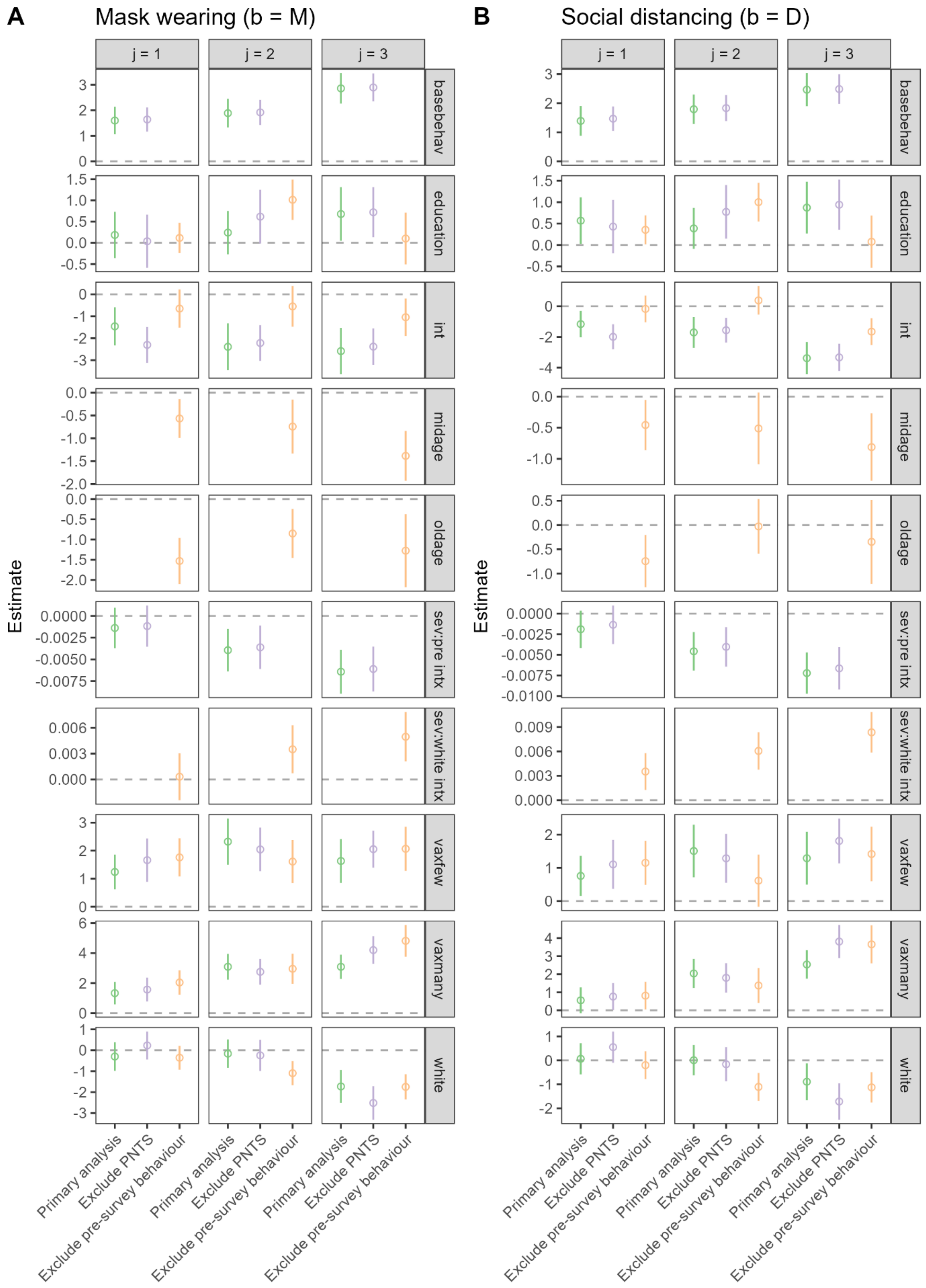


***Figure S5**. Estimated model coefficients for individual-level covariates, comparing the primary analysis (green) and sensitivity analyses excluding individuals responding prefer not to say (PNTS; purple) or excluding data on participant behaviour in the month prior to the survey (orange). Points*

*and error bars represent, respectively, means and 95% confidence intervals estimated from robust standard errors.*

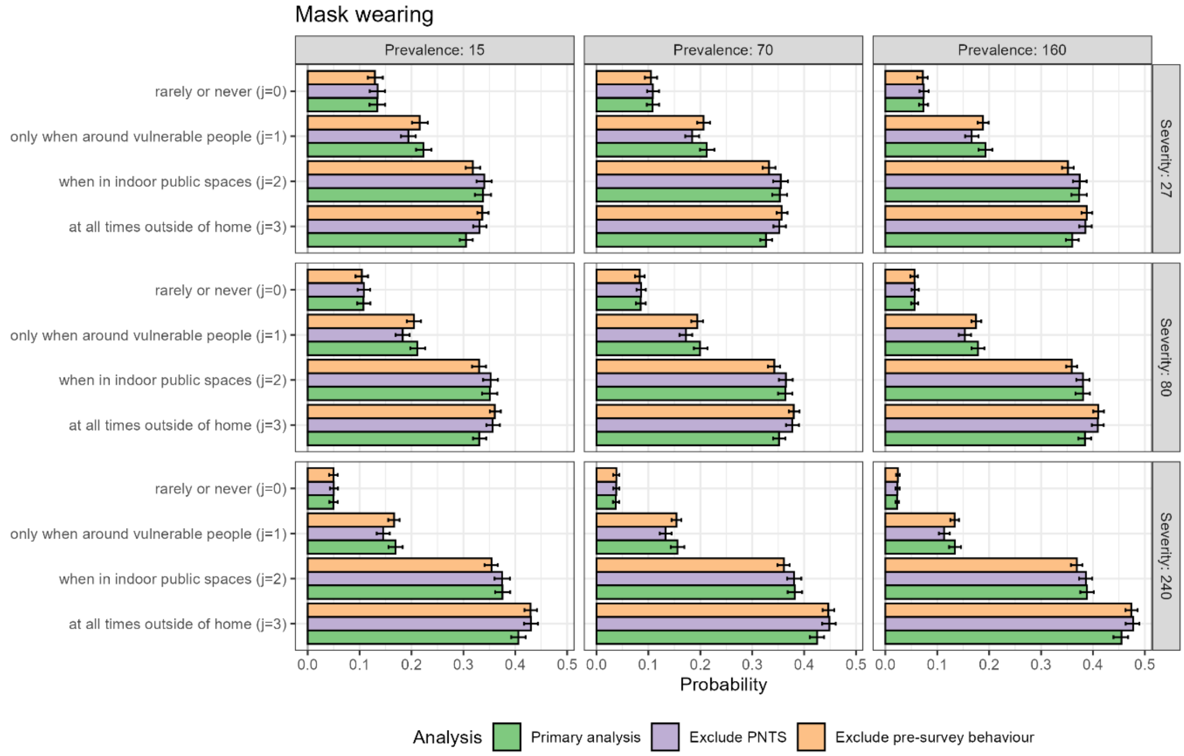


***Figure S6***. *Predicted probabilities of facemask wearing choices among the sample population when varying severity (rows) and prevalence (columns), comparing the primary analysis (green) to sensitivity analyses (purple, orange). These results assume low severity uncertainty, low prevalence uncertainty and the communication of risk only including text. Bar lengths and error bars represent, respectively, means and 95% uncertainty intervals. PNTS = prefer not to say.*

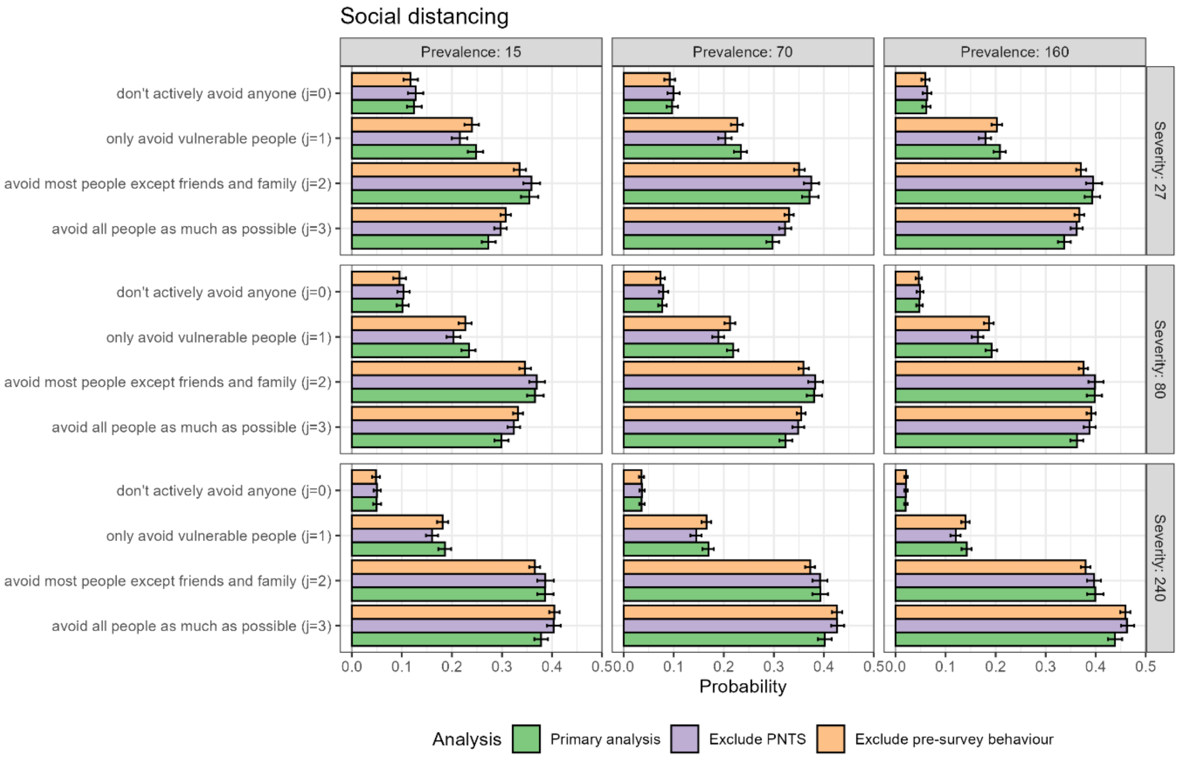


***Figure S7.*** *Predicted probabilities of social distancing choices among the sample population when varying severity (rows) and prevalence (columns), comparing the primary analysis (green) to sensitivity analyses (purple, orange). These results assume low severity uncertainty, low prevalence uncertainty and the communication of risk only including text. Bar lengths and error bars represent, respectively, means and 95% uncertainty intervals. PNTS = prefer not to say.*

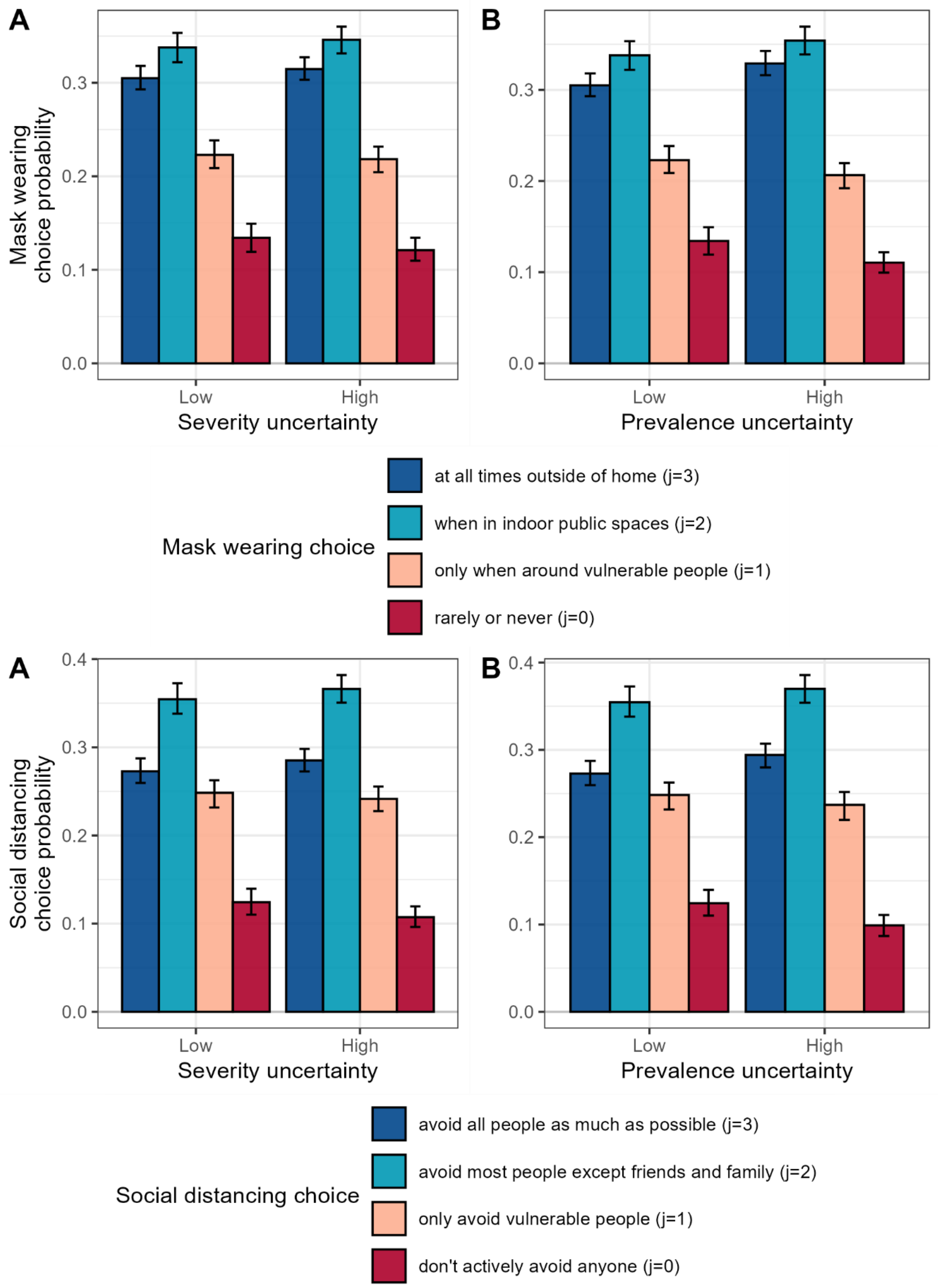


***Figure S8***. *Predicted probabilities of behavioural choices among the sample population when varying severity uncertainty and prevalence uncertainty, given lowest severity and lowest prevalence levels. The reference case here includes low severity uncertainty, low prevalence uncertainty and the communication of risk only including text. Bar heights and error bars represent, respectively, means and 95% uncertainty intervals.*

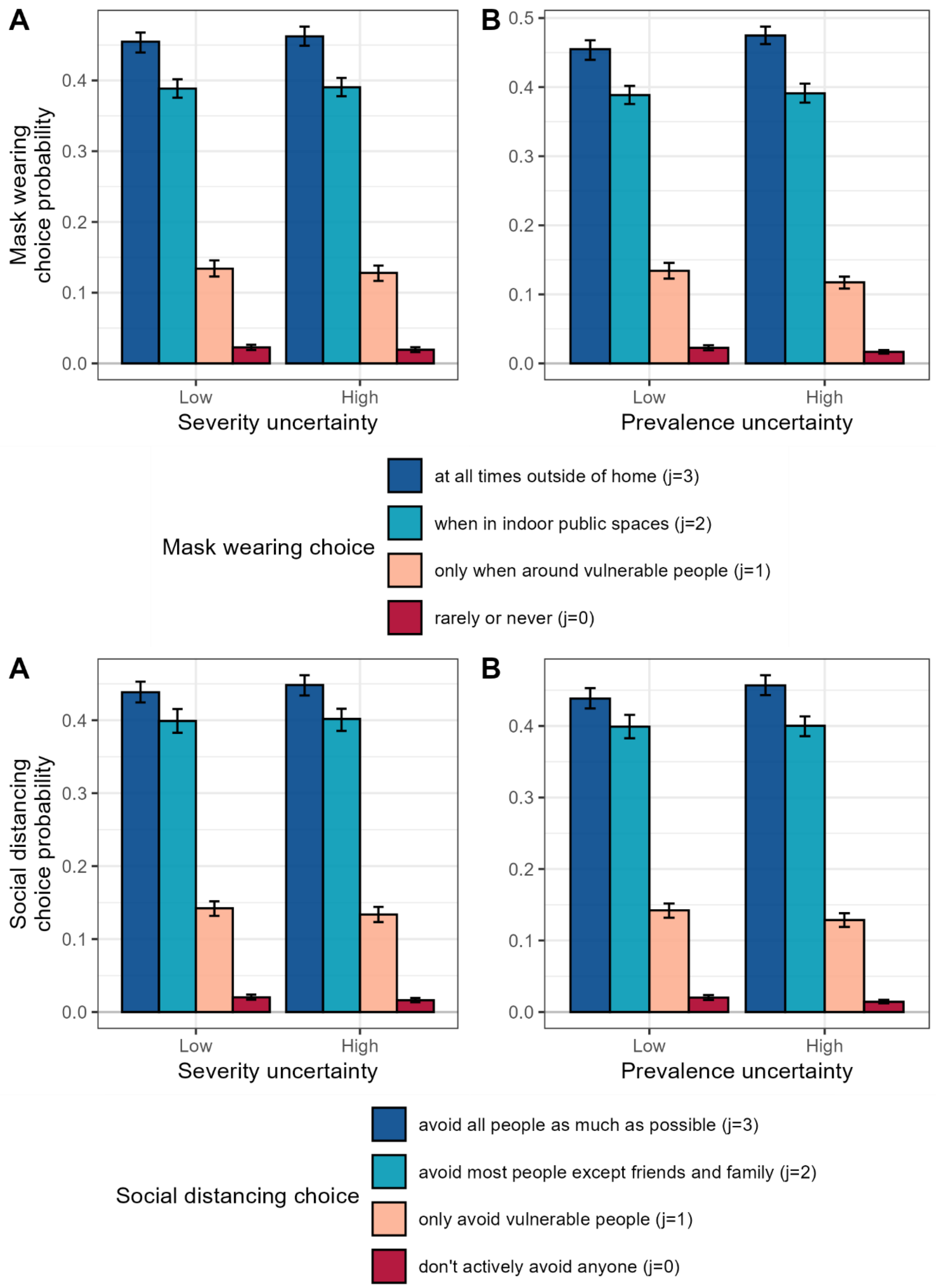


***Figure S9***. *Predicted probabilities of behavioural choices among the sample population when varying severity uncertainty and prevalence uncertainty, given highest severity and highest prevalence levels. The reference case here includes low severity uncertainty, low prevalence uncertainty and the communication of risk only including text. Bar heights and error bars represent, respectively, means and 95% uncertainty intervals.*
